# Antiviral efficacy of oral ensitrelvir versus oral ritonavir-boosted nirmatrelvir in COVID-19

**DOI:** 10.1101/2025.05.18.25327861

**Authors:** William HK Schilling, Podjanee Jittamala, Phrutsamon Wongnak, James A Watson, Simon Boyd, Viravarn Luvira, Tanaya Siripoon, Thundon Ngamprasertchai, Elizabeth M Batty, Ellen Beer, Shivani Singh, Tanatchakorn Asawasriworanan, Timothy Seers, Koukeo Phommasone, Terry John Evans, Varaporn Kruabkontho, Thatsanun Ngernseng, Jaruwan Tubprasert, Mohammad Yazid Abdad, Wanassanan Madmanee, Jindarat Kouhathong, Kanokon Suwannasin, Watcharee Pagornrat, Tianrat Piteekan, Borimas Hanboonkunupakarn, Kittiyod Poovorawan, Manus Potaporn, Attasit Srisubat, Bootsakorn Loharjun, Kesinee Chotivanich, Mallika Imwong, Sasithon Pukrittayakamee, Arjen M Dondorp, Nicholas PJ Day, Watcharapong Piyaphanee, Weerapong Phumratanaprapin, Nicholas J White, the PLATCOV Collaborative Group

## Abstract

**Background:** Ensitrelvir is an oral antiviral treatment for COVID-19 with the same molecular target as ritonavir-boosted nirmatrelvir - the current oral first-line treatment. There have been no direct comparisons between the two drugs.

**Methods:** In an open label controlled adaptive pharmacometric platform trial, low-risk adult patients aged 18-60 years with early symptomatic COVID-19 (<4 days of symptoms) were randomised concurrently to one of eight treatment arms including ensitrelvir, ritonavir-boosted nirmatrelvir, and no study drug. The primary endpoint was the rate of oropharyngeal viral clearance assessed in a modified intention-to-treat population (mITT), defined as patients with ≥3 days of follow-up. Viral clearance rate was derived under a Bayesian hierarchical linear model fitted to the log_10_ viral densities in standardised duplicate oropharyngeal swab eluates taken daily over five days (14 measurements). This trial is registered at ClinicalTrials.gov (NCT05041907).

**Findings:** Between March 2023 and April 2024 the three study arms randomised 604 patients concurrently in Thailand and Lao PDR (ensitrelvir 202; ritonavir-boosted nirmatrelvir 207; no study drug 195) among 903 patients enrolled. All patients recovered uneventfully. Ensitrelvir was very well tolerated and did not cause dysgeusia. Median (interquartile range) estimated SARS-CoV-2 clearance half-lives were 5.9 hours (4.0 to 8.6) with ensitrelvir; 5.2 hours (3.8 to 6.6) with nirmatrelvir; and 11.6 hours (8.1 to 14.5) with no study drug. Viral clearance following ensitrelvir was 82% (95% credible interval, CrI: 61 to 104%) faster than no study drug and 16% (95% CrI: 5 to 25%) slower than ritonavir-boosted nirmatrelvir. Viral rebound occurred in 15 (7%) of the nirmatrelvir group and 10 (5%) of the ensitrelvir group (p=0.4).

**Conclusions:** Both ensitrelvir and nirmatrelvir markedly accelerate oropharyngeal SARS-CoV-2 viral clearance. Ensitrelvir is an efficacious and well tolerated alternative to currently available antivirals in treating COVID-19.

**Funding:** “Finding treatments for COVID-19: A phase 2 multi-centre adaptive platform trial to assess antiviral pharmacodynamics in early symptomatic COVID-19 (PLAT-COV)” is supported by the Wellcome Trust Grant ref: 223195/Z/21/Z through the COVID-19 Therapeutics Accelerator.

**Research in context:** *Evidence before this study:* We searched PubMed for studies published in English from Jan 1, 2020, to April 10, 2025, using the terms: “randomised” AND [“nirmatrelvir OR paxlovid”] AND “ensitrelvir”. Both ritonavir-boosted nirmatrelvir and ensitrelvir have shown in-vivo antiviral activity and clinical benefit, but there have been no direct randomised head-to-head comparisons. Comparisons between the preregistration studies are confounded by substantial differences in the study populations, and timing of the studies.

*Added value of this study:* Comparison of antiviral drug efficacy using clinical endpoints is difficult-‘hard endpoints’ such as hospitalisation or death require prohibitively large sample sizes due to their rarity, and classification of more frequently encountered milder symptoms are imprecise. By contrast, this pharmacometric approach provides a quantitative measure of antiviral effects in patients with tractable sample sizes. This randomised study provides the first direct comparison of the in-vivo antiviral effects of ritonavir-boosted nirmatrelvir and ensitrelvir. Both drugs markedly accelerate SARS-CoV-2 viral clearance. An individual patient meta-analysis of all drugs included in the study confirms these drugs to have the most potent anti-SARS-CoV-2 antiviral effect.

*Implications of all the available evidence:* Both ritonavir-boosted nirmatrelvir and ensitrelvir have potent in-vivo antiviral activity in patients with early COVID-19. Ensitrelvir can be considered an efficacious and well-tolerated alternative to currently available antivirals. Candidate antivirals and antiviral combinations for respiratory viruses (including COVID-19 and Influenza) should be assessed and compared using this method.

## Introduction

Coronavirus disease 2019 (COVID-19) remains prevalent throughout the world and, although it has become an increasingly mild illness for most people as population immunity has increased and viral virulence has decreased, it still causes significant morbidity in immunocompromised and elderly patients. Two effective oral antiviral drugs (nirmatrelvir and molnupiravir) are currently available. Ritonavir-boosted nirmatrelvir is the more potent, but it is very expensive, commonly causes troubling dysgeusia (“bad taste”), and is associated with a long list of potential drug-drug interactions and there are concerns over symptomatic viral rebound after stopping the medication. There is very limited availability outside high-income settings (1). Molnupiravir is well tolerated but less potent (2), and this has led some countries not to adopt it in their guidelines. There are also concerns that it might generate more pathogenic or drug-resistant mutant viruses (2, 3, 4). These perceived drawbacks, costs and a lack of available alternatives have limited the use of oral antiviral drugs in COVID-19.

Ensitrelvir, like nirmatrelvir, is a 3C-like SARS CoV2 main protease inhibitor (M^pro^), with the potential advantages of increased stability and slower elimination (5). It can be given once daily (nirmatrelvir requires twice daily dosing) and does not require ritonavir boosting. Ensitrelvir is registered in Japan and Singapore and has been given to more than 1 million people, but it has not been compared directly with other antiviral drugs. The increasing rarity of hospitalisation and death in COVID-19, in marked contrast to five years ago, means that prohibitively large comparative studies in high-risk groups are now needed to detect clinically important differences between antiviral drugs. However, acceleration in viral clearance reflects clinical benefit in COVID-19 (6, 7, 8). We present a head-to-head randomised controlled platform trial comparison of the *in vivo* antiviral activities of ensitrelvir versus ritonavir-boosted nirmatrelvir in adults with early symptomatic COVID-19.

## Methods

PLATCOV is an ongoing phase 2 open label, multi-centre, randomised, controlled adaptive pharmacometric platform trial currently running in Thailand, Brazil, Nepal and Lao PDR (ClinicalTrials.gov: NCT05041907) (9). The trial provides a standardised quantitative comparative method for the *in vivo* assessment of potential antiviral treatments in low-risk adults with early symptomatic COVID-19. Potential antiviral treatments enter the randomised trial when they become available and leave when pre-specified endpoints are reached. The platform trial began recruitment on 30 September 2021. The initial drugs studied were ivermectin, favipiravir, remdesivir and the casivirimab/imdevimab monoclonal antibody cocktail (9, 10, 11, 12). All these arms reached the prespecified endpoints for efficacy or lack of efficacy and have now stopped. Additional arms were introduced subsequently (molnupiravir, ritonavir-boosted nirmatrelvir, fluoxetine, tixagevimab/cilgavimab monoclonal antibody cocktail, the combination treatment of molnupiravir plus ritonavir-boosted nirmatrelvir, and hydroxychloroquine) (2, 13). The primary outcome measure is the rate of viral clearance estimated under a linear model fitted to the log_10_ oropharyngeal viral densities over the first five days following randomisation in comparison with first the negative control (no treatment) and then the positive control (ritonavir-boosted nirmatrelvir). Originally the trial evaluated viral clearance over 7 days, but the marked shortening of natural viral clearance in recent years has meant that evaluation over a shorter period now has greater discriminative value (14). The change from seven to five days evaluation occurred on 5 March 2024 based on unblinded data from other treatment arms not including the blinded ensitrelvir (14). The evaluation of ensitrelvir was conducted in the Hospital for Tropical Diseases, Faculty of Tropical Medicine, Mahidol University, Bangkok, Thailand and Mahosot hospital, Vientiane, Lao PDR. After a detailed explanation of study procedures and requirements all patients provided fully informed written consent. PLATCOV is coordinated and monitored by the Mahidol Oxford Tropical Medicine Research Unit (MORU) in Bangkok. The trial was overseen by a trial steering committee (TSC), was conducted according to Good Clinical Practice principles and was approved by the two local IRB/ECs and the Oxford Tropical Research Ethics Committee. Its results were reviewed regularly by a data and safety monitoring board (DSMB). The ongoing platform trial is registered at ClinicalTrials.gov (NCT05041907).

### Randomisation and interventions

Block randomisation was performed for each site via a centralised web-app designed by MORU software engineers using RShiny®, hosted on a MORU webserver. At enrolment, after obtaining fully informed consent and entering the patient details, the app provided the study drug allocation. The “no study drug” arm comprised a minimum proportion of 20% of patients at all times, with uniform randomisation ratios applied across the active treatment arms. All patients received standard symptomatic treatment. The ensetrelvir comparative analysis includes only patients from Thailand and Lao PDR enrolled between the 17^th^ of March 2023 and 21st April 2024 (as the test drugs were unavailable at the other study sites). Oral ensitrelvir (Xocova™: Shionogi & Co., Ltd.), and ritonavir-boosted nirmatrelvir (Paxlovid™: Pfizer) were given in standard doses (appendix page 9). A loading dose of 375mg ensitrelvir (three tablets) was given on the first day and 125mg (one tablet) was given daily for the next four days. Nirmatrelvir 300mg with 100mg ritonavir (separate tablets) was given twice daily for five days. During this period, patients were also randomised to tixagevimab/cilgavimab, fluoxetine, hydroxychloroquine, ritonavir-boosted nirmatrelvir + molnupiravir combination, and nitazoxanide.

### Participants and procedures

Previously healthy adults aged between 18 and 60 years were eligible for trial enrolment if they understood the study procedures and requirements and gave fully informed consent for participation, reported symptoms of COVID-19 for less than 4 days (<96hrs), were SARS-CoV-2 positive (defined either as a nasal lateral flow antigen test which became positive within two minutes (STANDARD™ Q COVID-19 Ag Test, SD Biosensor, Suwon-si, Korea), or a positive PCR test with a cycle threshold value (Ct) <25 [all viral gene targets]) within the previous 24hrs), had oxygen saturation ≥96% measured by pulse oximetry, were unimpeded in activities of daily living, and agreed to adhere to all procedures, including availability and contact information for follow-up visits. Exclusion criteria included taking any concomitant medications or drugs, chronic illness or condition requiring long-term treatment or other significant comorbidity, laboratory abnormalities discovered at screening (haemoglobin <8g/dL, platelet count <50,000/uL, abnormal liver function tests, eGFR <70 mls/min/1.73m^2^), pregnancy (a urinary pregnancy test was performed in females), or actively trying to become pregnant, lactation, or contraindication or known hypersensitivity to any of the proposed therapeutics, currently participating in a COVID-19 therapeutic or vaccine trial or evidence of pneumonia (although imaging was not required).

Enrolled patients were admitted to the study ward or managed as outpatients, as per patient preference (none of the admissions were for clinical reasons, but for ease of adherence with the study procedures, or for self-isolation). All treatments were directly observed or video recorded. After randomisation and baseline procedures (appendix page 7) oropharyngeal swabs (two swabs from each tonsil) were taken as follows. A flocked swab (COPAN FLOQSwabs^®^) was rotated against the tonsil through 360° four times and placed in COPAN URT™ viral transport medium (3mL). Swabs were transferred at 4-8°C, aliquoted, and then frozen at −80°C within 48hrs. Separate swabs from each tonsil were taken once daily from day 0 to day 5, then on days 6, 7, 10 and day 14. Each swab was processed and tested separately. Vital signs were recorded three times daily by the patient (initial vital signs on the first day were recorded by the study team), and symptoms and any adverse effects were recorded daily.

The TaqCheck™ SARS-CoV-2 Fast PCR Assay (Applied Biosystems^®^, Thermo Fisher Scientific, Waltham, Massachusetts) quantitated viral loads (expressed as RNA copies per mL). This multiplexed real-time PCR method detects the SARS-CoV-2 N and S genes, and human RNase P gene in a single reaction. RNase P was used in the linear model to adjust for variation in sample human cell content (see Statistical Analysis Plan [SAP]). Viral loads were quantified against ATCC^®^ heat-inactivated SARS-CoV-2 (VR-1986HK™ strain 2019-nCoV/USA-WA1/2020) standards. The laboratory team was blinded to treatment allocation and the clinical investigators were blinded to the virology results until the study arm was terminated. Whole genome sequencing was performed to identify viral variants and allocate genotypes (appendix pages 15-16). Adverse events were graded according to the Common Terminology Criteria for Adverse Events v.5.0 (CTCAE). Summaries were generated if the adverse event was ≥grade 3 and was new, or had increased in intensity. Serious adverse events were recorded separately and reported to the DSMB.

### Outcome measures and statistical analysis

The primary outcome measure was the rate of viral clearance. This was expressed as a slope coefficient and estimated under a Bayesian hierarchical linear model with random effect terms for the individual patient slope and intercept (9). The model was fitted to the daily log_10_ viral load measurements between days 0 and 5 (14 measurements per patient), using weakly informative priors and treating non-detectable viral loads (CT value=40) as left-censored (appendix pages 10-11) (15). The treatment effect was defined as the multiplicative change (%) in the viral clearance rate, relative to the no study drug arm (15). The viral clearance rate (i.e. slope coefficient from the model fit) can also be expressed as a clearance half-life (t_1/2_ = log_10_ 0.5/slope). Thus a 50% increase in viral clearance rate equals a 33% reduction in clearance half-life. Secondary outcomes were: all cause hospitalisation for clinical deterioration (until day 28); time to fever clearance up to day 7; time to symptom resolution up to day 7; and viral rebound. Patients were defined as febrile at baseline if ≥ one axillary temperature measurement within 24 hours of randomisation was ≥37.5°C. Resolution of fever was defined as an axillary temperature ≤37.0°C for at least 24 hours. Resolution of symptoms was defined as no reported symptoms. Times to resolution of fever and symptoms were assessed using survival methods as the data were right-censored at the last visit, and described using restricted mean survival time. Comparisons between different treatment arms used the log-rank test, while the relative change in the rate of time to fever/symptom resolution was estimated from the Cox proportional hazards model. Viral rebound was defined as a mean daily oropharyngeal eluate viral load which declined to <100 genomes/mL for ≥2 consecutive days then rose to >1000 genomes/mL at any time thereafter. Proportions were compared using Pearson’s chi-square test. For each studied intervention in the PLATCOV trial the sample size was adaptive based on prespecified futility and success stopping rules (appendix page 12).

Full details of the trial procedures have been published previously (2, 10, 12, 16). The comparison with the most effective drug (currently nirmatrelvir) terminates when the intervention is shown to be inferior, non-inferior, or superior to the positive control arm using a 10% non-inferiority margin (Figure S1 appendix page 12). If a stopping rule was not met after 200 patients had been enrolled and evaluated, the arm was stopped anyway. All stopping decisions were prespecified and made using data from contemporaneously randomised patients only. Apart from the trial statisticians (JAW, PW), the clinical investigators were all blinded to the qPCR results, and the laboratory was blinded to the treatment allocations.

All analyses were done in a modified intention-to-treat (mITT) population, comprising patients who had >2 days follow-up data. A sensitivity analysis was performed using a non-linear model fitted to the serial viral densities, which allows for an initial increase in densities followed by a log-linear decrease (exact specification is given in the appendix pages 10-11). All models included site and calendar time as a covariate for the slope and intercept. For the meta-analysis of all small molecule antiviral interventions in the study we included random effects on the slope term by time period, breaking the whole study period into 10 bins with approximately equal numbers of patients in each time period. This allowed adjustment for temporal trends confounding comparison of interventions which were not assessed concurrently. Model fits were compared using approximate leave-one-out comparison as implemented in the package *loo*. All data analysis was done in R version 4.3.2. Posterior distributions were approximated using Hamiltonian Monte Carlo in *stan* via the *rstan* interface (17). 4000 iterations were run over 4 independent chains with 2000 iterations for burn-in. Convergence was assessed visually from the trace plots (Figure S2 appendix page 18) using the R-hat statistic (a value <1.1 was considered acceptable convergence) (18). Goodness of fit was assessed by plotting the residuals over time and comparing the daily median model predictions with the observed values (Figure S3 appendix page 19). All point estimates are given with 95% credible intervals, defined by the 2.5% and the 97.5% quantiles of the posterior distribution. All code and data are openly accessible via a GitHub repository: https://github.com/jwatowatson/PLATCOV-Ensitrelvir.

## Results

The ensitrelvir arm started enrolment on 17th March 2023. By that time ritonavir-boosted nirmatrelvir and no study drug were the positive and negative controls respectively. Initially, the pre-specified interim analyses compared ensitrelvir to the concurrent no study drug arm to assess antiviral efficacy. The first interim analysis, using data from 27 patients randomised to ensitrelvir and 29 concurrent negative controls, demonstrated that ensitrelvir had already met the criteria for efficacy (probability >0.9 of >20% increase in viral clearance) so, without interruption, ensitrelvir then entered a non-inferiority assessment with the “positive control” ritonavir-boosted nirmatrelvir. In April 2024, the pre-specified maximum number of participants had been recruited and evaluated although inferiority/non-inferiority thresholds had not been met. By then 202 patients had been randomised concurrently to ensitrelvir, 207 to ritonavir-boosted nirmatrelvir, and 195 to no study drug. 15 patients were excluded from the analyses because they withdrew from the study, resulting in a mITT population of 589 included into the non-inferiority assessment and subsequent analyses (Figure 1). Of these, 577 (98%) patients were from Thailand, and 12 (2%) were from Laos (Table 1). 545 (93%) had received at least one COVID-19 vaccine dose. The mean (SD) interval from symptom onset to randomisation was 1.8 (0.8) days and the geometric mean baseline viral density in oropharyngeal eluates was 5.1 log_10_ genomes per mL (SD 1.4). The baseline viral loads in the ensitrelvir arm were slightly lower (0.3 log_10_ genomes per mL) than in the ritonavir-boosted nirmatrelvir and no study drug arm. The model takes into account starting viral loads through random effects.

**Figure 1:**
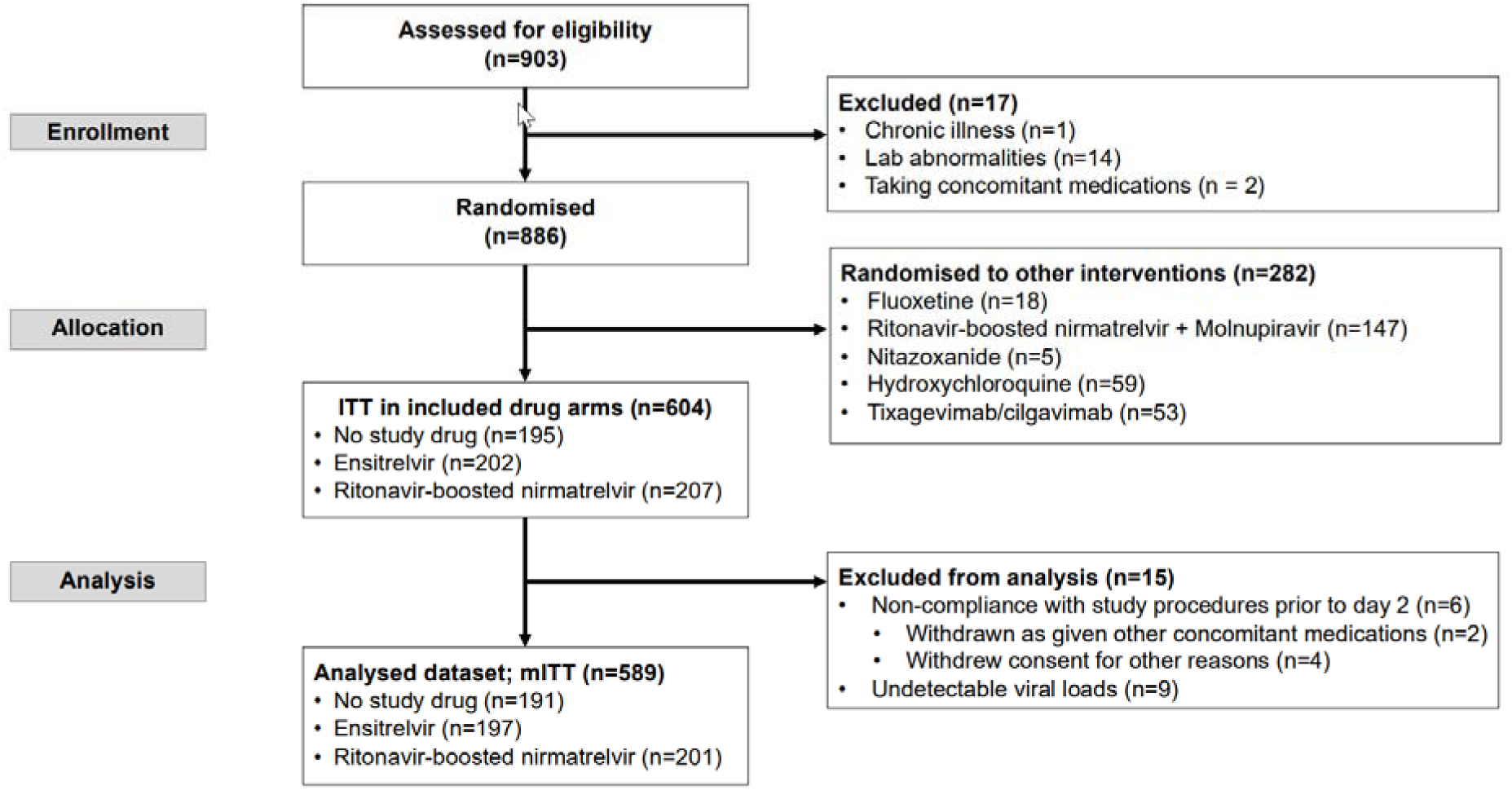
Study CONSORT diagram for the ensitrelvir versus ritonavir-boosted nirmatrelvir versus no study drug analysis. ITT, modified intention-to-treat. mITT, modified intention-to-treat.

**Table 1:** Baseline patient characteristics. Data are n (%) or mean (SD).

|  | Ritonavir-boosted<br>nirmatrelvir<br>(n = 201) | Ensitrelvir<br>(n = 197) | No study drug<br>(n=191) |
| --- | --- | --- | --- |
| Site: |  |  |  |
| • Laos | 4 (2) | 4 (2) | 4 (2) |
| • Thailand | 197 (98) | 193 (98) | 187 (98) |
| Sex: |  |  |  |
| • Male | 53 (26) | 61 (31) | 58 (30) |
| • Female | 148 (74) | 136 (69) | 133 (70) |
| Age (years) | 33.0 (9.4) | 32.5 (8.9) | 34.1 (9.3) |
| BMI (kg/m<sup>2</sup>) | 23.3 (4.7) | 23.3 (4.7) | 24.0 (4.2) |
| Weight (kg) | 61.8 (14.5) | 62.5 (15.8) | 63.9 (13.1) |
| Symptom duration (days) | 1.8 (0.8) | 1.8 (0.9) | 1.8 (0.9) |
| Baseline viral densities<br>(log<sub>10</sub> genomes/mL) | 5.2 (1.4) | 4.9 (1.4) | 5.2 (1.4) |
| Vaccinated | 188 (94) | 177 (90) | 180 (94) |

### Virological responses

Both ensitrelvir and ritonavir-boosted nirmatrelvir accelerated viral clearance. By day 3 the median viral densities were approximately 100-fold lower in both ensitrelvir and ritonavir-boosted nirmatrelvir arms compared to patients receiving no study drug, (Figure 2). Under a linear model fitted to all viral load data up to day 5, the rates of viral clearance were 82% (95% credible interval, CrI: 61 to 104%) faster with ensitrelvir and 116% (95% CrI: 91 to 142%) faster with nirmatrelvir relative to the no study drug arm (Figure S4, appendix page 20). The median estimated viral clearance half-lives under the linear model were 5.2 hours (IQR: 3.8 to 6.6) with nirmatrelvir; 5.9 hours (IQR: 4.0 to 8.6) with ensitrelvir; and 11.6 hours (IQR: 8.1 to 14.5) in the contemporaneous no study drug arm. I.e. median virus clearance half-life was approximately halved by both ensitrelvir and nirmatrelvir (Figure 3a). In the non-inferiority comparison, viral clearance was 16% (95% CrI: 5 to 25%) slower with ensitrelvir relative to nirmatrelvir (probability less than the non-inferiority margin of 10%: 0.86) (Figure 3b).

**Figure 2:**
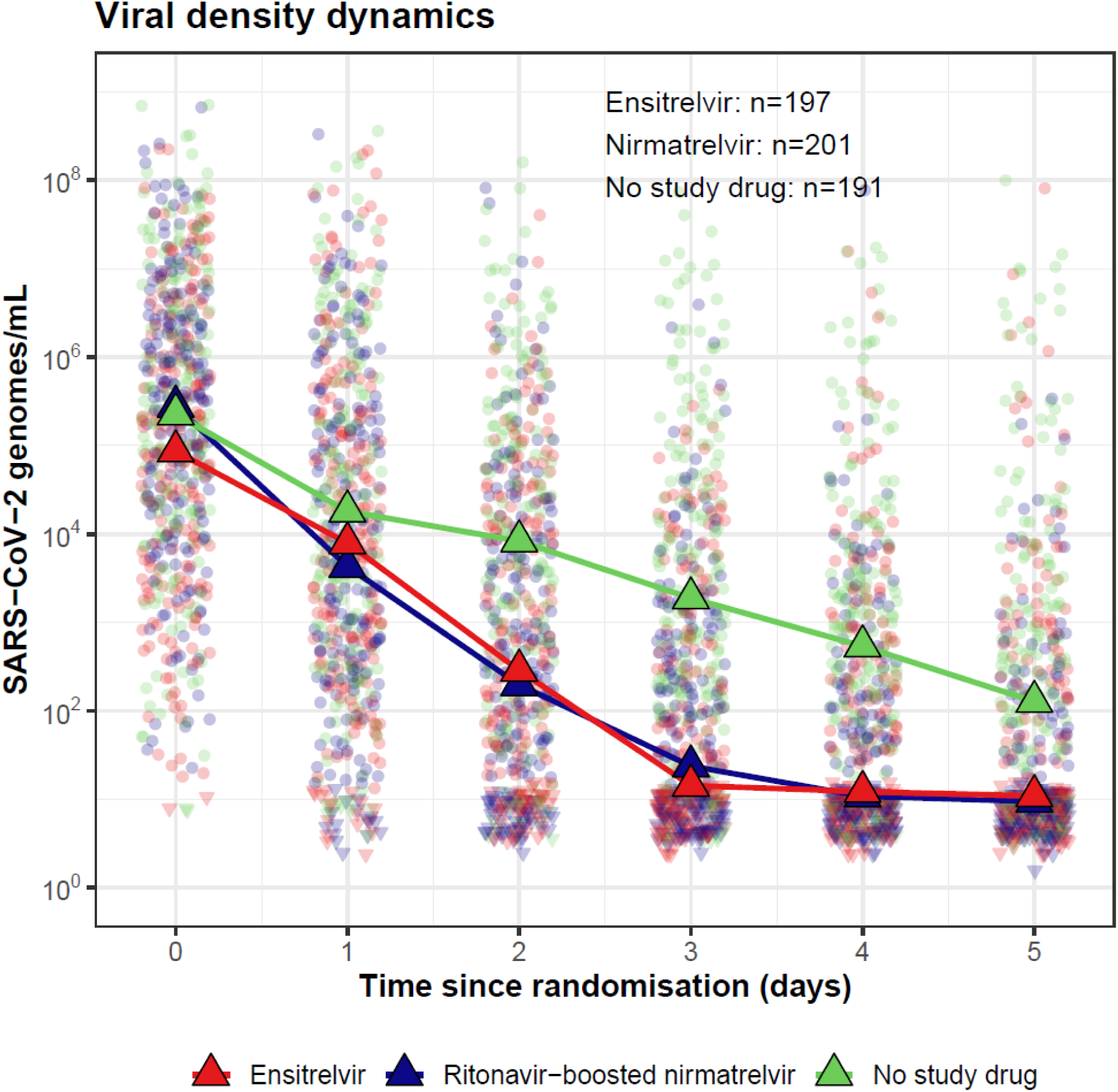
SARS-CoV-2 oropharyngeal median viral loads over time in the three contemporaneous randomised arms (individual data points shown as circles).

**Figure 3:**
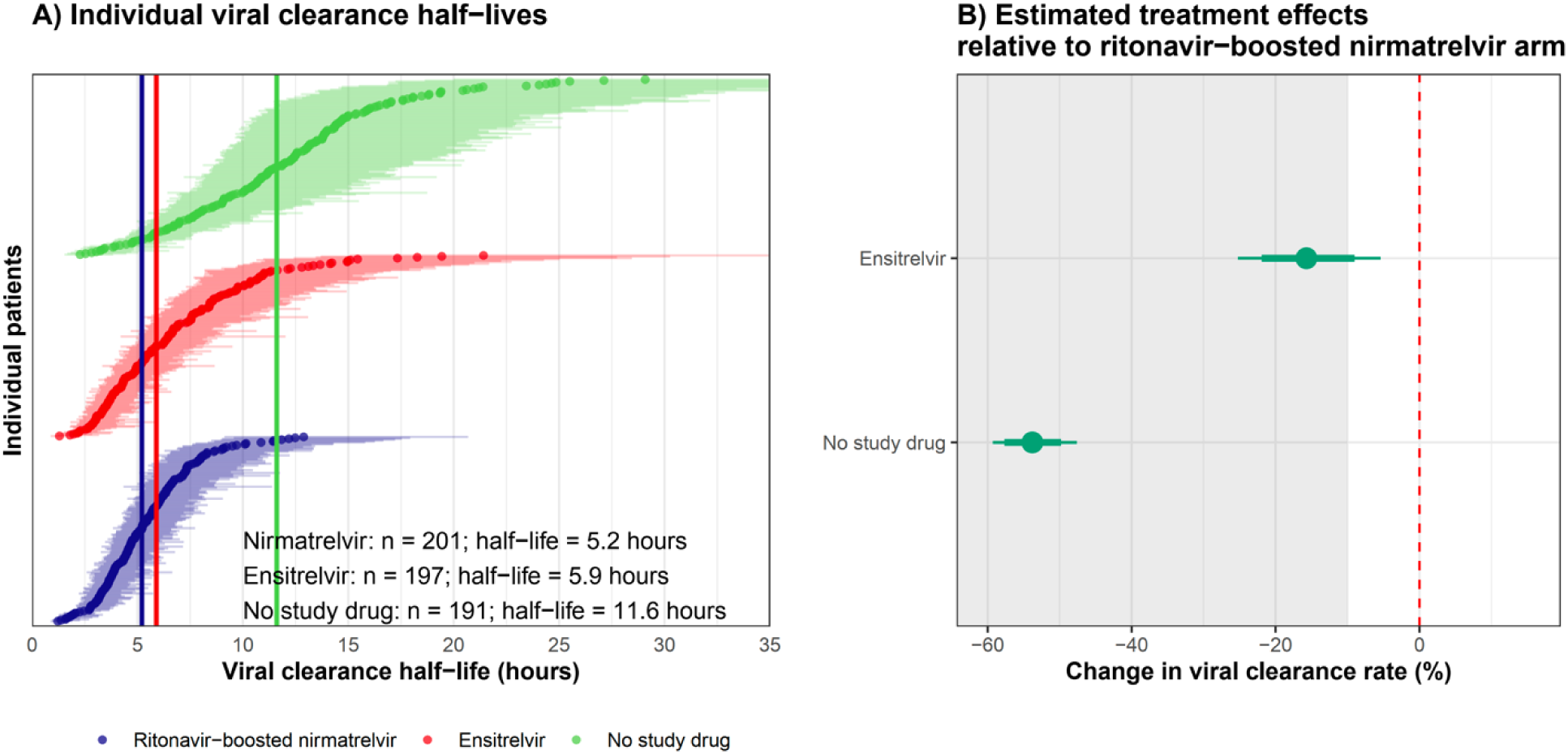
Left panel: Individual patient estimated virus clearance half-lives grouped by treatment arm (point estimates and 80% credible intervals are shown). The vertical dashed lines show the median half-lives in each group. Right panel: The estimated treatment effects relative to nirmatrelvir under the linear model (the grey zone shows the inferiority zone relative to nirmatrelvir). Thick and thin error bars indicate the 80% and 95% credible intervals, respectively.

### Clinical responses

No patients developed severe disease although seven patients were admitted to hospital-ensitrelvir (n=2), nirmatrelvir (n=3) and no study drug (n=2). Two patients reported fatigue related to COVID-19 (nirmatrelvir n=1; no study drug n=1) and one had a likely drug-interaction (nirmatrelvir arm). The other hospitalisations were considered to be unrelated to COVID-19 or the study medications (Table S2, appendix page 16).

The mean duration of fever was 1.4 days (95% CI: 1.2 to 1.7 days) in the no study drug arm, and 1.2 days (95% CI: 0.9 to 1.6 days) in both the ensitrelvir and ritonavir-boosted nirmatrelvir arms. The time to fever clearance was not significantly different between patients who received either ensitrelvir or ritonavir-boosted nirmatrelvir and those in the no study drug arm.

The restricted mean duration of symptoms over one week was 5.4 days (95% CI: 5.2 to 5.7 days) following ensitrelvir, 5.4 days (95% CI: 5.1 to 5.6 days) following ritonavir-boosted nirmatrelvir; and 5.9 days (95% CI: 5.6 to 6.1 days) following no study drug. Relative to the no study drug arm, symptom resolution was 32% faster in the ensitrelvir arm (95% CI: 3% slower to 78% faster) and 38% faster in the ritonavir-boosted nirmatrelvir arm (95% CI: 3% faster to 86% faster) (Figure 4B).

**Figure 4:**
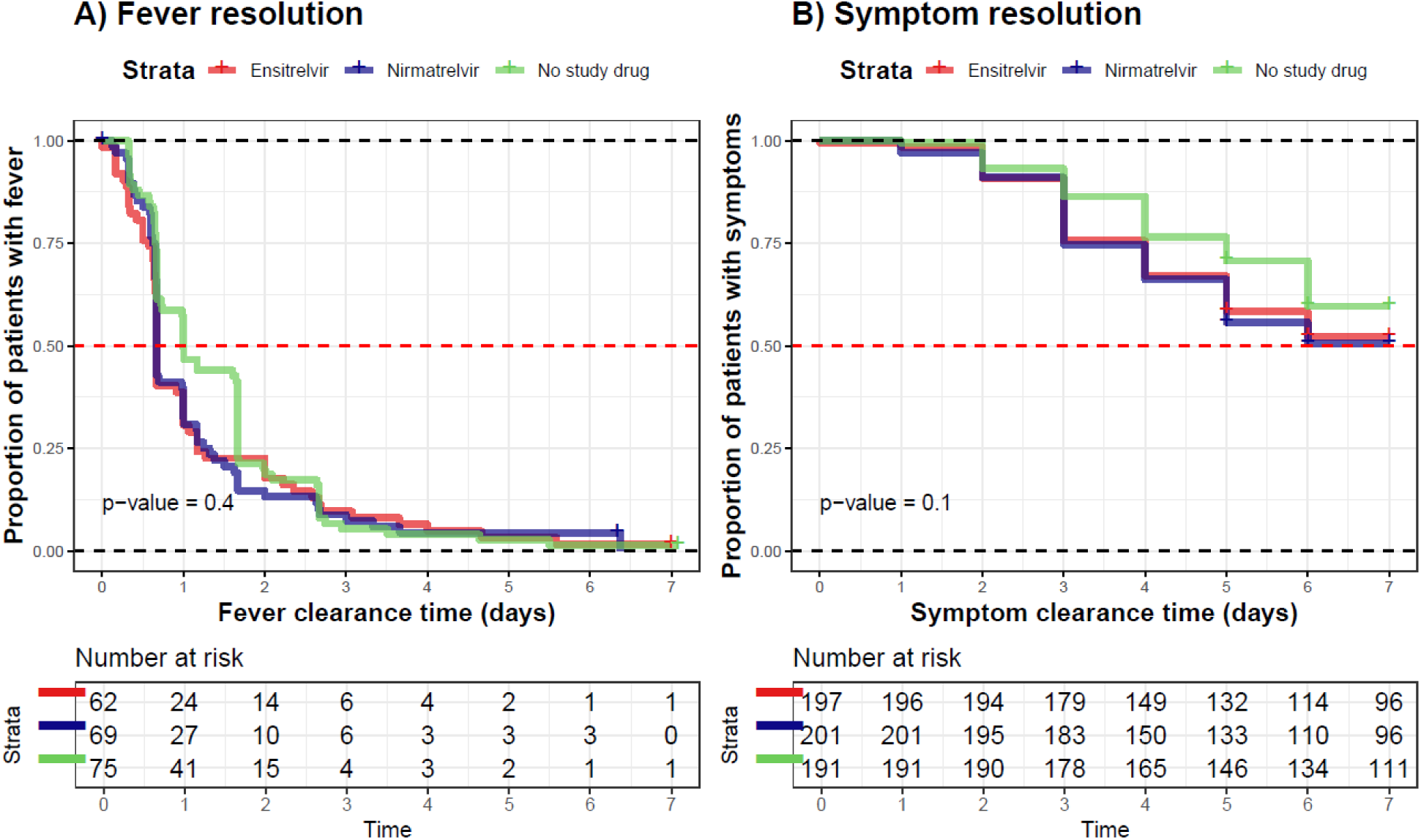
Kaplan-Meier curve for the time to fever resolution (left) and the time to symptom resolution (right) 7-day post-randomisation across three treatment groups, with the p-value of the log-rank test indicated.

### Tolerability and adverse effects

The oropharyngeal swabbing procedure and all treatments were well-tolerated. Two patients in the ritonavir-boosted nirmatrelvir group discontinued their treatment because of interactions with their concomitant drugs (Figure 1). 52 (26%) out of 201 patients in the ritonavir-boosted nirmatrelvir group, 2 (1%) out of 197 patients in the ensitrelvir group, and 1 (1%) out of 191 patients in the no study drug group complained of dysgeusia (experiencing bitter/metallic taste) (Figure S5, appendix page 21).

### Viral rebound

The proportion of patients with viral rebound was similar among the different groups; p-value = 0.6); 7% (15/201) for nirmatrelvir, 5% (10/197) for ensitrelvir, and 7% (13/191) for no study drug (Figure S6, appendix page 22), but the median (IQR) time interval to viral rebound was longer for nirmatrelvir; 9.9 days (IQR: 6.8 to 13.8) than for ensitrelvir 5.9 days (IQR: 5.0 to 6.7) and for no study drug 6.9 days (IQR: 6.0 to 9.0).

### Individual patient data meta-analysis

To compare antiviral effects of all the unblinded small molecule drugs tested in the PLATCOV platform trial, we performed an individual patient data meta-analysis using patients recruited in Thailand and Lao PDR. This comprised recipients of ivermectin (9), remdesivir (10), favipiravir (12), fluoxetine (19), molnupiravir (2), nirmatrelvir (2), ensitrelvir, or no study drug (hydroxychloroquine, nitazoxanide, molnupiravir + ritonavir-boosted nirmatrelvir remain blinded). The analysis population comprised 1,157 patients randomised between 30 September 2021 and 22 April 2024 in Thailand and Laos, with a total of 16,171 qPCR measurements (82% were above the lower limit of quantification). As the interventions were not randomised concurrently, and thus temporal confounding is expected, the analysis adjusted for calendar time (14) Figure S7, appendix page 23). The no study drug arm spanned the entire study period.

Under the linear model the two interventions reported previously to have no clinical antiviral effect, ivermectin and favipiravir (9, 12), had very similar virus clearance rates to the no study drug arm (Figure 5). Remdesivir, previously reported to have a moderate effect on viral clearance (10), was estimated to increase viral clearance relative to no study drug by 44% (95% CI: 16 to 77%). The increase viral clearance with molnupiravir was estimated to be 60% (95% CrI: 34 to 92%), and the increase with fluoxetine was estimated to be 20% (95% CrI: 3 to 39%).

**Figure 5:**
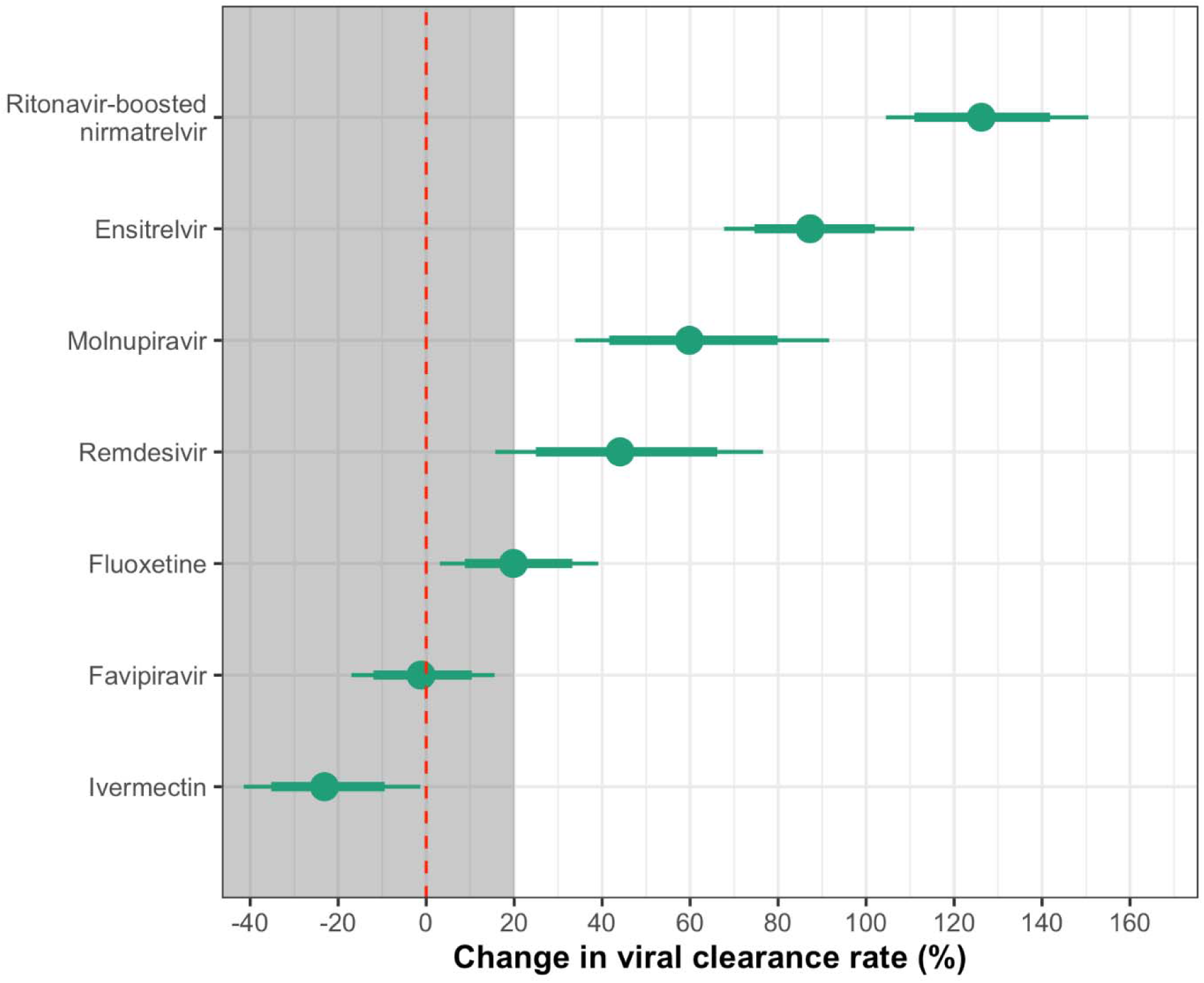
Estimated treatment effects relative to no study drug arm from the individual patient data meta-analysis under the linear model (the grey zone shows the futility zone). Thick and thin error bars indicate the 80% and 95% credible intervals, respectively.

In the overall comparison, ensitrelvir and nirmatrelvir accelerated viral clearance relative to no study drug by 87% (95% CrI: 68 to 111%) and 126% (95% CrI: 105 to 151%), respectively.

## Discussion

This first comparative *in vivo* pharmacodynamic assessment of ensitrelvir and ritonavir-boosted nirmatrelvir confirms that ensitrelvir has potent antiviral activity in treating COVID-19. This effect was slightly less than with ritonavir-boosted nirmatrelvir, although it did not reach the prespecified statistical threshold for non-inferiority. The meta-analysis of this platform trial indicated that ensitrelvir was more potent than all the other antivirals that have been evaluated other than ritonavir-boosted nirmatrelvir.

The main indication for oral antiviral treatment in COVID-19 is to prevent disease progression. Early in the pandemic this was measured by reduction in hospitalisation or death (20, 21, 22, 23). Now the disease is milder, antiviral efficacy is increasingly assessed by reduction in symptom severity and duration, and the presence of longer-term sequelae. Although COVID-19 can still be a significant illness in the frail or immunocompromised, increasing population immunity and the decreasing virulence of the evolving viral variants mean that COVID-19 in the general population is often either asymptomatic or results in a mild self-limiting upper respiratory tract infection. This makes the evaluation of antiviral drugs difficult. Two almost identical studies of ritonavir-boosted nirmatrelvir were conducted two years apart (with no appreciable loss of antiviral activity of the drug). The first study early in the pandemic demonstrated a clear acceleration in symptom resolution and a life-saving benefit, but the second larger study, conducted when the disease had become milder, struggled to show significant benefit in symptom resolution (21, 24). Similarly, SCORPIO-HR, a double-blind, randomised, placebo-controlled trial comparison of ensitrelvir to placebo conducted in the Omicron era in a predominantly vaccinated population, did not meet its primary endpoint of a statistically significant reduction in the time to sustained symptom resolution.

Both ritonavir boosted nirmatrelvir and ensetrelvir were generally well tolerated although over 25% of the patients who received ritonavir-boosted nirmatrelvir actively complained of dysgeusia. There were no clear differences in clinical responses and, although there slightly fewer viral rebounds in the ensitrelvir arm, the overall number of rebounds was very low.

Pharmacometric studies assessing rates of viral clearance measure antiviral efficacy *in vivo* and can be used to compare treatments efficiently with smaller numbers than conventional Phase III studies (6–8). They provide a solution to the current difficulties in assessing antivirals in COVID-19 and comparing efficacy between those assessed in earlier epochs (when the disease was much more severe). Until now, the limited choice of outpatient COVID-19 treatments has been dictated by availability, cost, tolerability, drug interactions, route of administration, and perceived drawbacks and benefits. To this can be added the difficulty in demonstrating comparable benefit to already licensed treatments in clinical trials, in order to satisfy the requirements for regulatory approval. For many governments, healthcare workers and individuals, antivirals for COVID-19 are no-longer considered necessary, as the disease is now generally mild and the drawbacks and costs are felt to outweigh the benefits. As a result, symptomatic individuals are often not treated for COVID-19. However, elderly, frail or immunocompromised patients do need antiviral treatment but many are also receiving other drugs which interact with ritonavir, and so cannot receive nirmatrelvir. Ensitrelvir has potent in vivo antiviral efficacy and it has the advantages of a lower pill burden. It also does not cause dysgeusia. Ensitrelvir does inhibit CYP3A4 (albeit to lesser extent than ritonavir) which means that many of the same concomitant medications are contraindicated. As ensitrelvir has the same molecular target and mechanism of action as nirmatrelvir there is overlap in resistance caused by target mutations, but these are very rare and they have not affected treatment efficacy.

Despite being the largest pharmacometrics assessment in COVID-19 this study has several limitations. We intentionally evaluated the interventions in low-risk adults with high viral burdens in order to optimise the comparative assessment of the different drugs, and not in high-risk patients or the elderly who are at greatest risk of disease progression. Protection against severe disease could not therefore be assessed in this study, and measures of clinical recovery would require a much larger study for confident assessment. Viral rebound was also a relatively rare event in the population studied in this trial. Finally, this comparison was conducted only in Southeast Asia. Population differences in immune status and pharmacokinetics may have affected therapeutic responses.

In summary, ensitrelvir has potent *in vivo* antiviral activity and was well-tolerated in the treatment of COVID-19. It provides some benefits over other currently available treatments. Affordable, efficacious, cost-effective and well-tolerated treatments are still needed for COVID-19.

## Supporting information

SAP

Protocol

## Data Availability

All data produced are available online at https://github.com/jwatowatson/PLATCOV-Ensitrelvir

## Acknowledgements

We thank all the patients with COVID-19 who volunteered to be part of the study. We thank the data safety and monitoring board (DSMB) (Tim Peto, André Siqueira, and Panisadee Avirutnan); the trial steering committee (TSC) (Nathalie Strub-Wourgaft, Martin Llewelyn, Deborah Waller, and Attavit Asavisanu); Sompob Saralamba and Tanaphum Wichaita for developing the RShiny randomisation app; and Mavuto Mukaka for invaluable statistical support. We also thank all the staff of the Clinical Trials Unit (CTU) at MORU, Thermo Fisher® for their excellent support with this project, and all the hospital staff at the Hospital for Tropical Diseases (HTD), as well as those involved in sample processing in MORU and the processing and analysis at the Faculty of Tropical Medicine, Mahidol University, molecular genetics laboratory and the malaria laboratory. We thank Shionogi & Co., Ltd. for the donation of ensitrelvir. We thank the Department of Medical Services, Ministry of Public Health, Thailand for the donation of the ritonavir-boosted nirmatrelvir (Paxlovid™). We thank the MORU Clinical Trials Support Group (CTSG) for data management, monitoring and logistics, and the purchasing, administration and support staff at MORU.

## Author’s contributions

WHKS-funding acquisition, investigation, methodology, project administration, supervision, validation and writing-original draft. PJ-investigation, methodology, project administration, supervision, validation and writing-original draft. PJ and WHKS contributed equally. JAW-conceptualisation, data curation, formal analysis, funding acquisition, methodology, visualisation and writing-original draft. SB-investigation, methodology, project administration, writing-original draft. VL, TS, TN-Investigation, methodology, supervision. EMB-data curation, formal analysis, visualisation. CC, JJC, SS, MS, VK, NT, JT-methodology, investigation, project administration. TN-data curation, software, supervision. WM, JK, KS, WP, NP-investigation, methodology. PH, BH, KP, VC-investigation, supervision. MP, AS, BL-resources. WRJT-methodology, supervision. KC, MI-formal analysis, investigation, resources, supervision. SP, AMD, MMT, WP, WP-methodology, investigation, resources, supervision. NPJD-funding acquisition, methodology, investigation, resources, supervision. NJW-conceptualisation, funding acquisition, methodology, supervision, validation and writing-original draft.

All Authors were involved in writing-review & editing.

JAW, WHKS, EMB, TN, MI and NJW have directly accessed and verified the underlying data reported in the manuscript.

## Data sharing statement

All code and de-identified participant data required for replication of the study’s endpoints are openly accessible via GitHub, as well as the study protocol and statistical analysis plan, from publication date onwards: https://github.com/jwatowatson/PLATCOV-Ensitrelvir. Individual Patient Data can be requested and may be shared according to the terms defined in the MORU data sharing policy with other researchers to use in the future from the date of publication. Further information on how to apply is found here: https://www.tropmedres.ac/units/moru-bangkok/bioethics-engagement/data-sharing.

