## Supplemental Data for "Antiviral efficacy of oral ensitrelvir versus oral ritonavir-boosted nirmatrelvir in COVID-19"

Supplementary materials

**Randomised comparison of the clinical antiviral efficacy of oral ensitrelvir versus oral ritonavir-boosted nirmatrelvir**

### List of Sites and Investigators (PLATCOV Collaborative Group)

**Sites**

1. Hospital for Tropical Diseases (HTD), Faculty of Tropical Medicine, Mahidol University, 420/6 Rajvithi Road, Bangkok, 10400, Thailand
2. Mahosot Hospital, Quai Fa Ngum, Vientiane, Laos

**Investigators**

Co-principal investigators:

Nicholas J White (1,2)

William Schilling (1,2)

Faculty of Tropical Medicine:

Site and Country Principal investigator:

Weerapong Phumratanaprapin^4^

Accountable Investigator:

Viravarn Luvira^4^

Study co-Investigators:

James Callery^1,2^

Nicholas PJ Day^1,2^

Sasithon Pukirttayakamee^1,4^

Phrutsamon Wongnak^1^

Simon Boyd^1,2^

Cintia Cruz^1,2^

Arjen M Dondorp^1,2^

James A Watson^1,2^

Watcharapong Piyaphanee^4^

Kittiyod Poovorawan^1,4^

Thundon Ngamprasertchai^4^

Tanaya Siripoon^4^

Borimas Hanboonkunupakarn^1,4^

Kesinee Chotivanich^1,2^

Podjanee Jittamala^1,3^

Mallika Imwong^1,5^

Janjira Thaipadungpanit^1,4^

Maneerat Ekkapongpisit^1^

Varaporn Kruabkontho^1^

Thatsanun Ngernseng^1^

Jaruwan Tubprasert^1^

Mohammad Yazid Abdad^1,2^

Elizabeth M Batty^1,2^

Shivani Singh ^1,2^

Tanatchakorn Asawasriworanan ^1^

Nuttakan Tanglakmankhong^1^

Wanassanan Madmanee^1^

Jindarat Kouhathong^1^

Kanokon Suwannasin^1^

Watcharee Pagornrat^1^

Nattaporn Piaraksa^1,4^

Ellen Beer ^1,2^

Bangplee Hospital:

Site Principal investigator:

Pongtorn Hanboonkunupakarn^6^

Co-investigator:

Sakol Sookprome^6^

Vajira Hospital:

Site Principal investigator:

Vasin Chotivanich^7^

Co-investigators:

Wiroj Ruksakul^7^

Chunlanee Sangketchon^8^

Brazil Site (Universidade Federal de Minas Gerais):

Site and Country Principal investigator:

Mauro M Teixeira^9^

Co-Investigators:

Pedro J Almeida^9^

Renato S Aguiar^10^

Franciele M Santos^10^

Laos Site – Mahosot Hospital

Elizabeth Ashley^2,11^

Manivanh Vongsouvath^11^

Koukeo Phommasone^11^

Audrey Dubot-Pérès^11^

Sisouphanh Vidhamaly^12^

Ammala Chingsanoon^12^

Danoy Chommanam^11^

Terry John Evans^11^

Vayouly Vidhamaly^11^

Latsaniphone Boutthasavong ^11^

Mayfong Mayxay^11^

Ministry of Public Health, Thailand

Manus Potaporn^13^

Attasit Srisubat^13^

Bootsakorn Loharjun^13^

1. Mahidol Oxford Tropical Medicine Research Unit, Faculty of Tropical Medicine, Mahidol University, Bangkok, Thailand

2. Centre for Tropical Medicine and Global Health, Nuffield Department of Medicine, Oxford University, Oxford, UK

3. Department of Tropical Hygiene, Faculty of Tropical Medicine, Mahidol University, Bangkok, Thailand

4. Department of Clinical Tropical Medicine, Faculty of Tropical Medicine, Mahidol University, Bangkok, Thailand

5. Department of Molecular Tropical Medicine and Genetics, Faculty of Tropical Medicine, Mahidol University, Bangkok, Thailand

6. Bangplee Hospital, Ministry of Public Health, Samut Prakarn province, Thailand

7. Faculty of Medicine, Navamindradhiraj University, Bangkok, Thailand

8. Faculty of Science and Health Technology, Navamindradhiraj University, Bangkok,Thailand

9. Clinical Research Unit, Center for Advanced and Innovative Therapies, Universidade Federal de Minas Gerais, Minas Gerais, Brazil

10. Department of Genetics, Ecology and Evolution, Institute of Biological Sciences, Universidade Federal de Minas Gerais, Minas Gerais, Brazil

11. Lao-Oxford-Mahosot Hospital-Wellcome Trust Research Unit, Mahosot Hospital, Vientiane, Lao PDR

12. Mahosot Hospital, Quai Fa Ngum, Vientiane, Lao PDR

13. Department of Medical Services, Ministry of Public Health, Nonthaburi, Thailand

### Ethics Approval

The trial was approved by local and national research ethics boards in Thailand (Faculty of Tropical Medicine Ethics Committee, Mahidol University, FTMEC Ref: TMEC 21-058, approval number MUTM 2021-057-03) and the Central Research Ethics Committee (CREC, Bangkok, Thailand, CREC Ref: CREC048/64BP-MED34), in Laos by the National Ethics Committee for Health Research (NECHR, Lao People’s Democratic Republic, Submission ID 2022.48) and the Federal Drug Administration (FDA, Lao People’s Democratic Republic, 13066/FDD_12Dec2022) and by the Oxford University Tropical Research Ethics Committee (OxTREC, Oxford, UK, OxTREC Ref: 24-21).

### Baseline procedures

Baseline investigations included a full clinical examination, blood sampling for hematology and biochemistry, an electrocardiogram and a chest radiograph (following local guidance in Thailand, but not a study requirement).

### Ensitrelvir additional information

Ensitrelvir (Shionogi Inc. Osaka, Japan) is given at a dose of three 125mg tablets (375mg) on day 1, then one 125mg tablet per day for a total treatment course of 5 days.

### Symptoms included in symptom questionnaire performed during study visits

- Fever
- Headache
- Dizziness
- Blurred vision
- Fatigue
- Cough
- Difficulty breathing
- Chest pain
- Running nose
- Loss of smell or taste
- Abdominal pain
- Loss of appetite
- Nausea
- Vomiting
- Diarrhoea
- Arthralgia
- Myalgia
- Itching
- Skin rash
- Sore throat
- Other (Free texts)

### Randomisation

The randomisation sheets were generated by the trial statistician. All new randomisation sheets and all updates of existing randomisation sheets were done using a pre-written R script which was stored on the randomisation Dropbox folder (owner is MORU, under custodianship of the head of MORU IT); this file is a full ‘Professional’ version with history recorded and only the trial statistician and head of IT had access. The file took the following inputs:

- Site codes (e.g. “th001”) for which to generate randomisation sheets;
- The set of arms available for randomisation in that site;
- The number of arm repeats per block (this is set to the minimum integer such that in each block there is an integer number for each arm);
- The randomisation data file from each site (which has the patient numbers for subjects already randomszed) named data-XXX.csv (where XXX is the site code), if this does not yet exist a blank .csv (headers only) is generated.

This R script is run every time a new site becomes active and every time the set of available arms changes. The output is a .csv file named rand-XXX.csv (where XXX is the site code). This overwrites the pre-existing file (which can be retrieved from the Dropbox version history). Each time the randomisation script is run, this is recorded on a log file*.*

The randomisation is done according to the following constraints:

- Blocks of 2*number of available arms;
- Additional ‘fuzziness’ by swapping one patient allocation per block at random (this can be swapped for any of the available arms) – this avoids knowing which arm the last patient per block will receive.

Each time an authorised member of the study team logs onto the web-app this is logged (timestamp and username).

Each time a new patient is randomised this is logged on to the file data-XXX.csv (where XXX is the site code) with the following information:

- Subject number
- Screening number
- Age
- Sex
- Member of study team username
- Timestamp

### Statistical Analysis

The original primary analysis consists of fitting Bayesian hierarchical (mixed effects) linear models to the serial log_10_ viral load data up until day 7. The marked shortening of natural viral clearance in recent years has meant that evaluation over a shorter period has greater discriminative value. The change from seven to 5 days evaluation occurred on 5 March 2024 based on unblinded data.^1^ All models encode residual error as a *t*-distribution with degrees of freedom estimated from the data. The *t*-distribution was chosen for robustness as the residual error is clearly non-Gaussian.^2,3^ The *t*-distribution error model also makes the inferences robust against model mis-specification (particularly for the linear models).^4^ All models include correlated individual random effect terms for both the intercept (baseline viral load) and the slope. All changes to the slope are defined as multiplicative changes on the log scale (i.e. a value of 0 equals no change).

The treatment effect is defined as the proportional change (expressed as a multiplicative term) in the population slope of the daily change in log_10_ viral load. The data are modelled on the log_10_ copies per mL scale, after conversion from Ct values using the standard curve generated from the 12 control concentrations (synthetic samples with known viral densities) from each 96 well plate. The standard curve transformation is done by fitting a linear mixed effects model (random slope and random intercept for each plate) to the control data: regressing the Ct values on the known log viral densities. This borrows information across plates and adjusts for batch effects.

For all models, we adjusted the intercept and slope for the study sites and calendar time. A subset of models also adjusted the slopes and intercepts for:

- Age
- Days since symptom onset
- Sex

All models adjust for human RNase P (proxy for the number of human cells in the sample). This is added as an independent linear predictor for each viral load measurement. We fitted the models using weakly informative priors (WIP) as prior distributions. The statistical analysis plan provides a detailed overview of the model structures <https://github.com/jwatowatson/PLATCOV-SAP/tree/main>.

***Analyses***

Platform trials can suffer from temporal confounding when interventions and controls are not all randomised concurrently. The primary three-way comparison between ensitrelvir, ritonavir-boosted nirmatrelvir, and no study drug only used concurrently randomised patients from one single centre. The meta-analysis of all unblinded small molecule drugs did not use concurrently randomised patients only. We included random effects on the slope term by time period, breaking the whole study period into 10 bins with approximately an equal number of patients in each time period. This allowed adjustment for temporal trends confounding comparison of interventions not assessed concurrently.

***Code and analysis plan***

All data, models and analytical output are on the linked GitHub repository: <https://github.com/jwatowatson/PLATCOV-Ensitrelvir>. This includes all data used in the analysis for full reproducibility of the results. The main markdown file (Ensitrelvir_analysis.qmd) runs the analysis for the primary three-way comparison. The markdown file Ensitrelvir_analysis_temporal_re.qmd runs the meta-analysis. The analyses for fever and symptom resolution are conducted in the fever_analysis.qmd and symptom_analysis.qmd files, respectively.

### Stopping rules

The stopping rules were determined using a simulation approach, based on previously modelled serial viral load data (3), such that approximately 50 patients are needed to demonstrate increases in the rate of viral clearance of ~50%, with control of both type 1 and type 2 errors at 10%. The prespecified decision criteria for stopping a treatment arm were either a model-based probability of <0.1 that the intervention did not accelerate viral clearance by >12.5% relative to no study drug (futility), or >0.9 that it did (success). In January 2023 this was changed to a 20% futility/success threshold. For the non-inferiority comparison, a 10% non-inferiority threshold was then chosen based on simulation analyses (see Statistical Analysis Plan for more details on the simulations). The updated diagram for how interventions can progress through the study is shown in Figure S1.

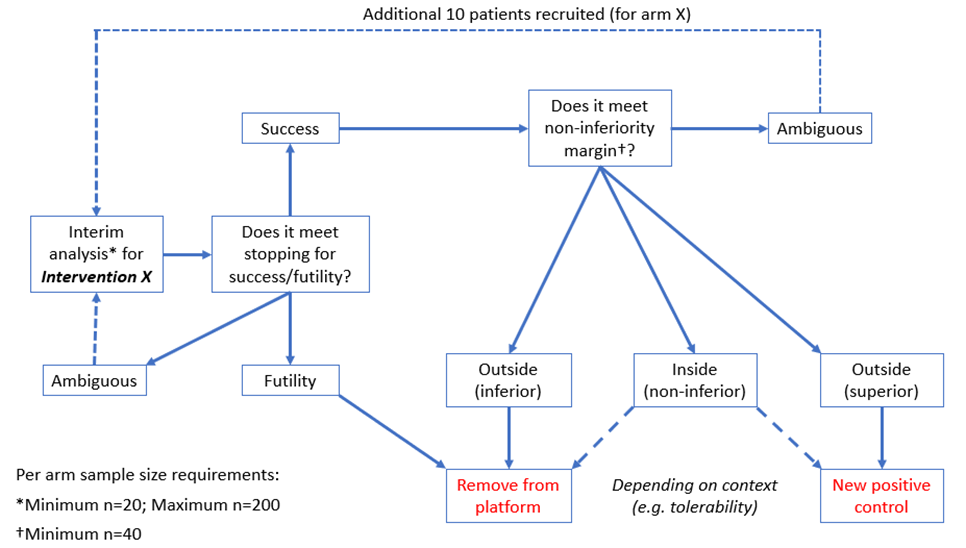

**Figure S1:** Schematic representation of the stopping rules as of January 2023.

### Adverse events (AE) for ensitrelvir and ritonavir-boosted nirmatrelvir

**Table S1:** Summary of adverse events (grade 3 and above) for ensitrelvir and ritonavir-boosted nirmatrelvir. *Patients were also classified as serious adverse events and are detailed in the serious adverse events table (Table S2). All AEs solicited were reported between days 0-7, day 14 and day 28.

|  | **All grades** | | | **Grade 3-4** | | |
| --- | --- | --- | --- | --- | --- | --- |
|  | Ensitrelvir (n = 197) | Nirmatrelvir (n = 201) | No drug (n = 191) | Ensitrelvir (n = 197) | Nirmatrelvir (n = 201) | No drug (n = 191) |
| Any adverse event (Grade ≥ 3) | 2 | 3 | 2 |  |  |  |
| Serious adverse event reported | 2 | 3 | 2 |  |  |  |
| Symptoms | | | | | | |
| Fever |  |  |  | 0 | 0 | 0 |
| Headache |  |  |  | 0 | 0 | 0 |
| Dizziness |  |  |  | 0 | 0 | 0 |
| Blurred vision |  |  |  | 0 | 0 | 0 |
| Fatigue |  |  |  | 0 | 1*** | 1*** |
| Cough |  |  |  | 0 | 0 | 0 |
| Difficulty breathing |  |  |  | 0 | 0 | 0 |
| Chest pain |  |  |  | 0 | 0 | 0 |
| Running nose |  |  |  | 0 | 0 | 0 |
| Loss of smell or taste |  |  |  | 0 | 0 | 0 |
| Abdominal pain |  |  |  | 0 | 0 | 0 |
| Loss of appetite |  |  |  | 0 | 0 | 0 |
| Nausea |  |  |  | 0 | 0 | 0 |
| Vomiting |  |  |  | 0 | 1*** | 0 |
| Diarrhoea |  |  |  | 0 | 0 | 1*** |
| Arthralgia |  |  |  | 0 | 0 | 0 |
| Myalgia |  |  |  | 0 | 0 | 0 |
| Itching |  |  |  | 0 | 0 | 0 |
| Skin rash |  |  |  | 0 | 0 | 0 |
| Other |  |  |  | 2*** | 1*** | 0 |
| Laboratory abnormalities | | | | | | |
| Creatinine |  |  |  | 0 | 0 | 0 |
| BUN |  |  |  | 0 | 0 | 0 |
| Sodium |  |  |  | 0 | 0 | 0 |
| eGFR |  |  |  | 0 | 0 | 0 |
| Potassium |  |  |  | 0 | 0 | 0 |
| ALT/SGPT |  |  |  | 0 | 0 | 0 |
| AST/SGOT |  |  |  | 0 | 0 | 0 |
| Total bilirubin |  |  |  | 0 | 0 | 0 |
| Direct bilirubin |  |  |  | 0 | 0 | 0 |
| Alkaline Phosphatase |  |  |  | 0 | 0 | 0 |
| LDH |  |  |  | 0 | 0 | 0 |
| Creatinine phosphokinase (CPK) |  |  |  | 0 | 0 | 0 |
| Anemia |  |  |  | 0 | 0 | 0 |
| Leukocytopenia |  |  |  | 0 | 0 | 0 |
| Neutropenia |  |  |  | 0 | 0 | 0 |
| Thrombocytopenia |  |  |  | 0 | 0 | 0 |

Serious Adverse Events

Table S2: Summary of Serious Adverse Events for ensitrelvir and ritonavir-boosted nirmatrelvir.

| **Number** | **Study arm** | **Final diagnosis** | **Relationship to trial drug** | **Resolved** |
| --- | --- | --- | --- | --- |
| 1 | Nirmatrelvir/ ritonavir | Bacterial tonsillitis^1^ | Not related | Yes |
| 2 | Nirmatrelvir/ ritonavir | COVID-19-related fatigue^2^ | Not related | Yes |
| 3 | Nirmatrelvir/ ritonavir | Nausea and Vomiting^3^ | Definitely related | Yes |
| 4 | No Study Drug | Acute diarrhoea^4^ | Not related | Yes |
| 5 | No Study Drug | COVID-19-related fatigue^5^ | Not related | Yes |
| 6 | Ensitrelvir | Motorcycle accident^6^ | Not related | Yes |
| 7 | Ensitrelvir | Motorcycle accident^7^ | Not related | Yes |

*^1^Participant presented with a 3-day history of high fever, sore throat and dysphagia. She was found to have acute bacterial tonsillitis and started on IV co-amoxiclav, PO levofloxacin and lidocaine for pain relief. She improved well on the ward with treatment and was discharged.*

*^2^Patient presented to the hospital with worsening fatigue, nausea and dizziness which had continued since the COVID-19 illness began. She was hypotensive (97/54) and tachycardic (HR 98) and had saturations of 96% on room air. She was treated with IV fluids and dimenhydrinate and improved on the ward. She was discharged without follow up.*

*^3^Patient was randomised to nirmatrelvir/ritonavir and after the second dose she self-administered one tablet of cafergot (which contains 1 MG of ergotamine tartrate and 100 MG of caffeine). Two hours later she experienced nausea and vomiting, numbness of both feet and hands, fatigue and difficulty breathing. The patient was hospitalised and had normal observations/examination and ECG. Nirmatrelvir/ritonavir was discontinued and patient was discharged without further follow up.*

*^4^Patient attended hospital due to acute diarrhoea and was treated with intravenous fluids. She was discharged the next day clinically resolved and required no further follow up.*

*^5^Patient experienced fatigue and dizziness whilst she was having her daily oropharyngeal swab on day 3. She had not been able to eat or drink for 1-2 days. She had 3 hours of observations and symptomatic supportive treatment but did not recover; she was therefore admitted into the hospital overnight. Her examination and observations were normal, urinary pregnancy test was negative, blood tests were not of clinical concern. She improved well on the ward and was discharged without follow up on*

*^6^Patient had motorcycle accident after having completed her medication course. There were no preceding symptoms or loss of consciousness. She was admitted into hospital overnight and discharged with a splint.*

*^7^Patient had a motorcycle accident with no preceding symptoms or loss of consciousness. She attended hospital for two nights and was diagnosed with abrasions of the left ankle, left knee and left elbow.*

### Supplementary Results

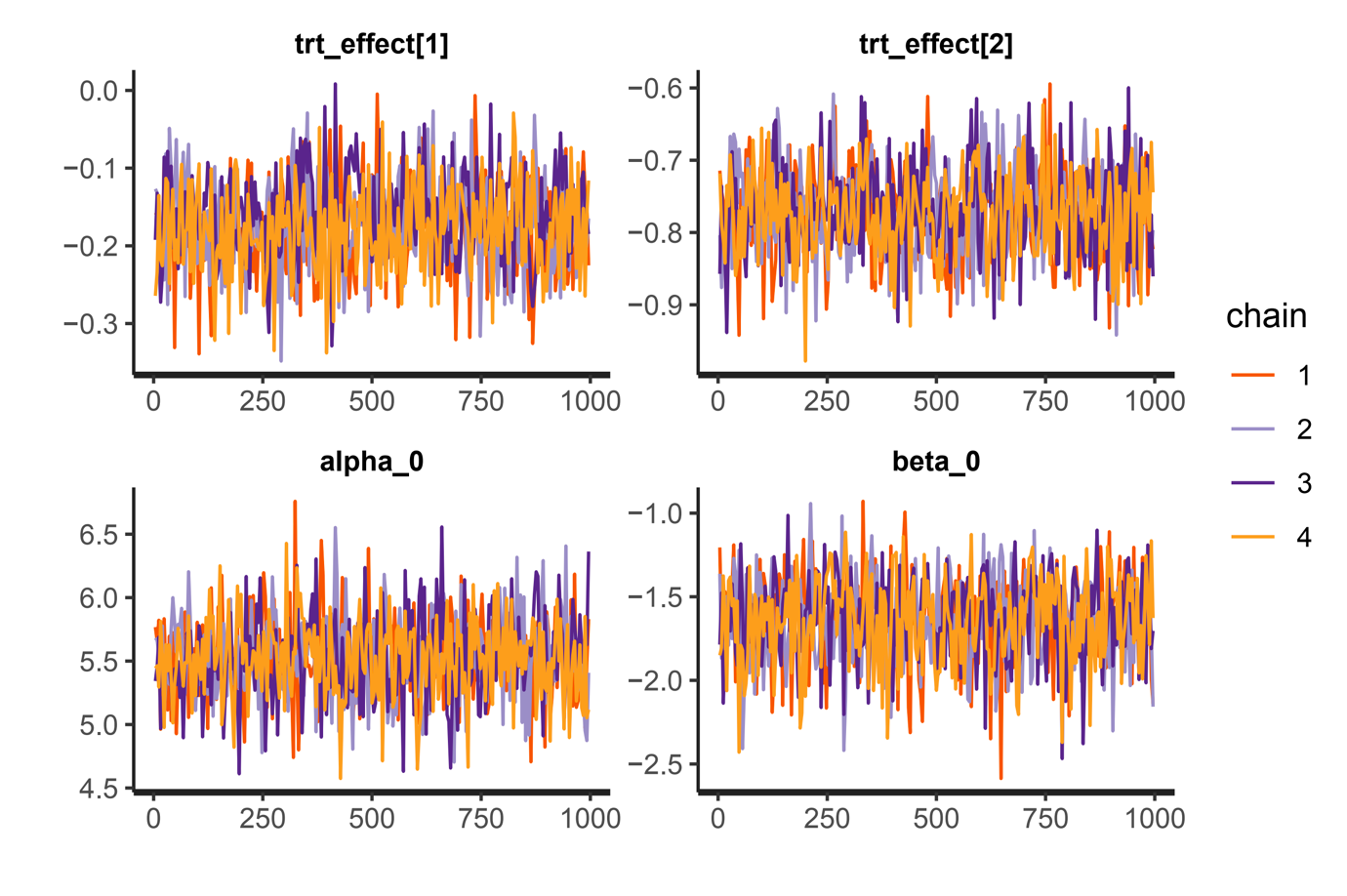

**Figure S2:** Traceplots indicating the convergence of model parameters from 4 MCMCs of the main analysis.

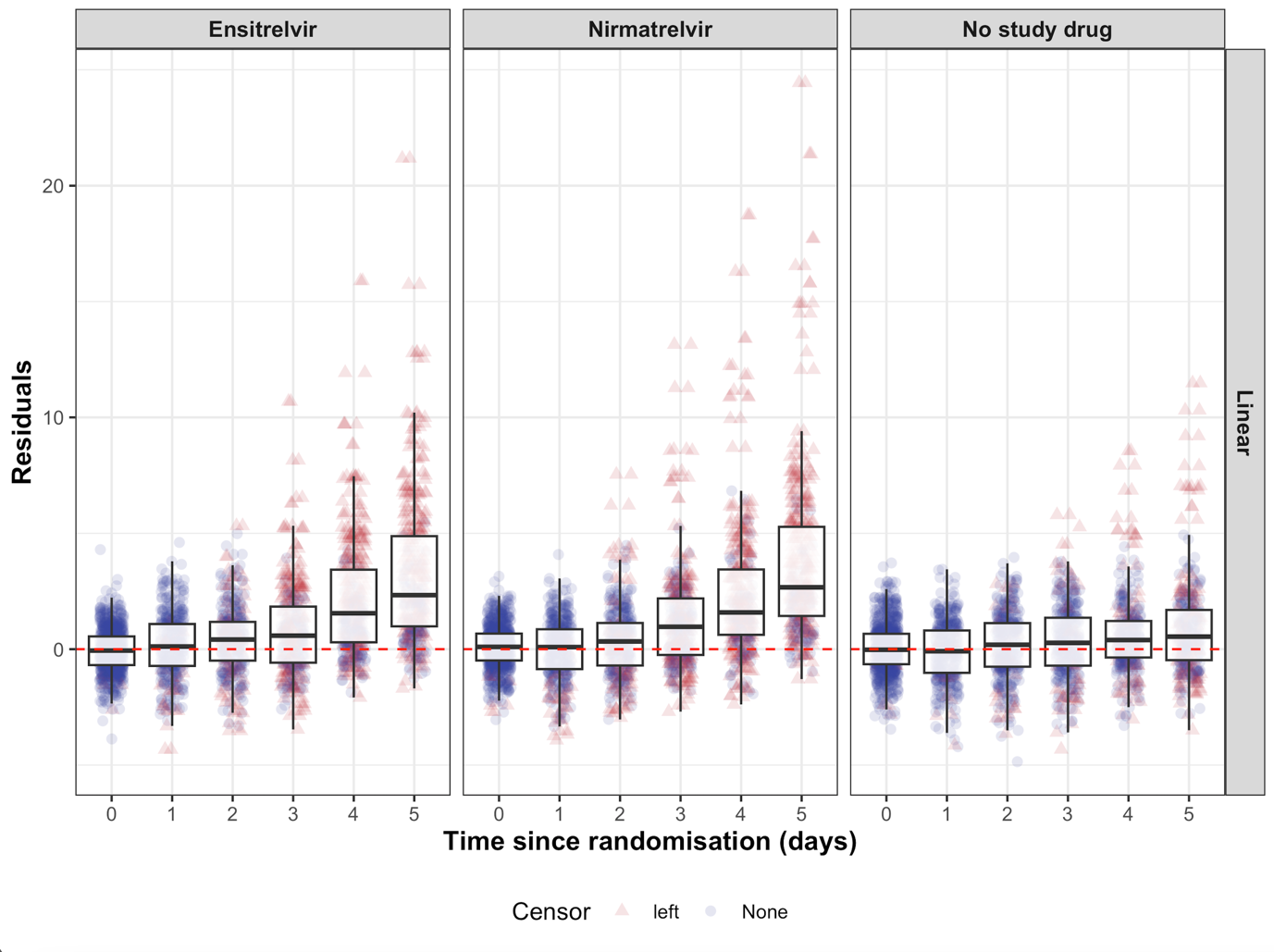
**Figure S3:** Residuals distribution over time of the main analysis

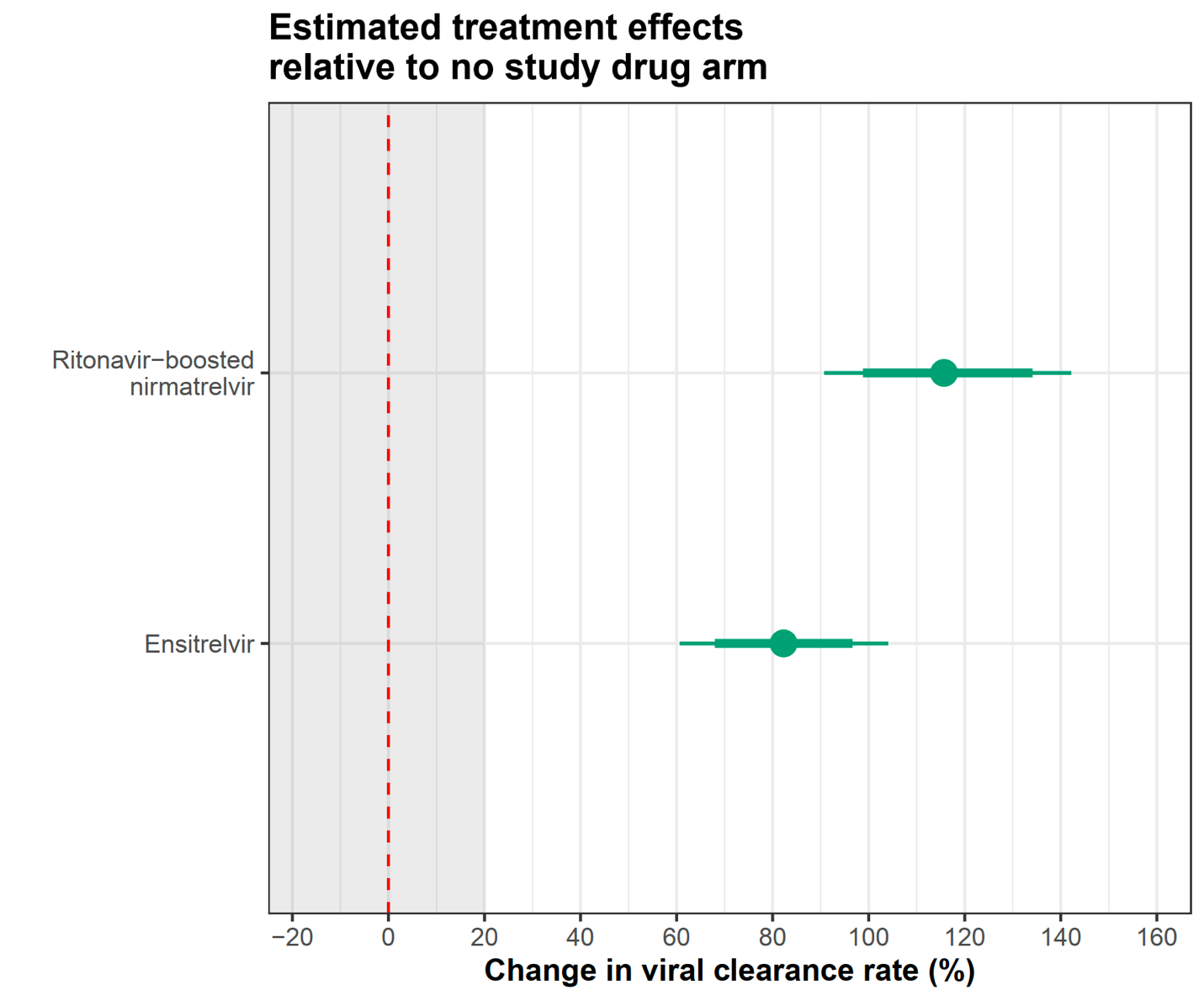

**Figure S4:** Estimated treatment effects relative to no study drug arm under the linear model (the grey zone shows the futility zone). Thick and thin error bars indicate the 80% and 95% credible intervals, respectively.

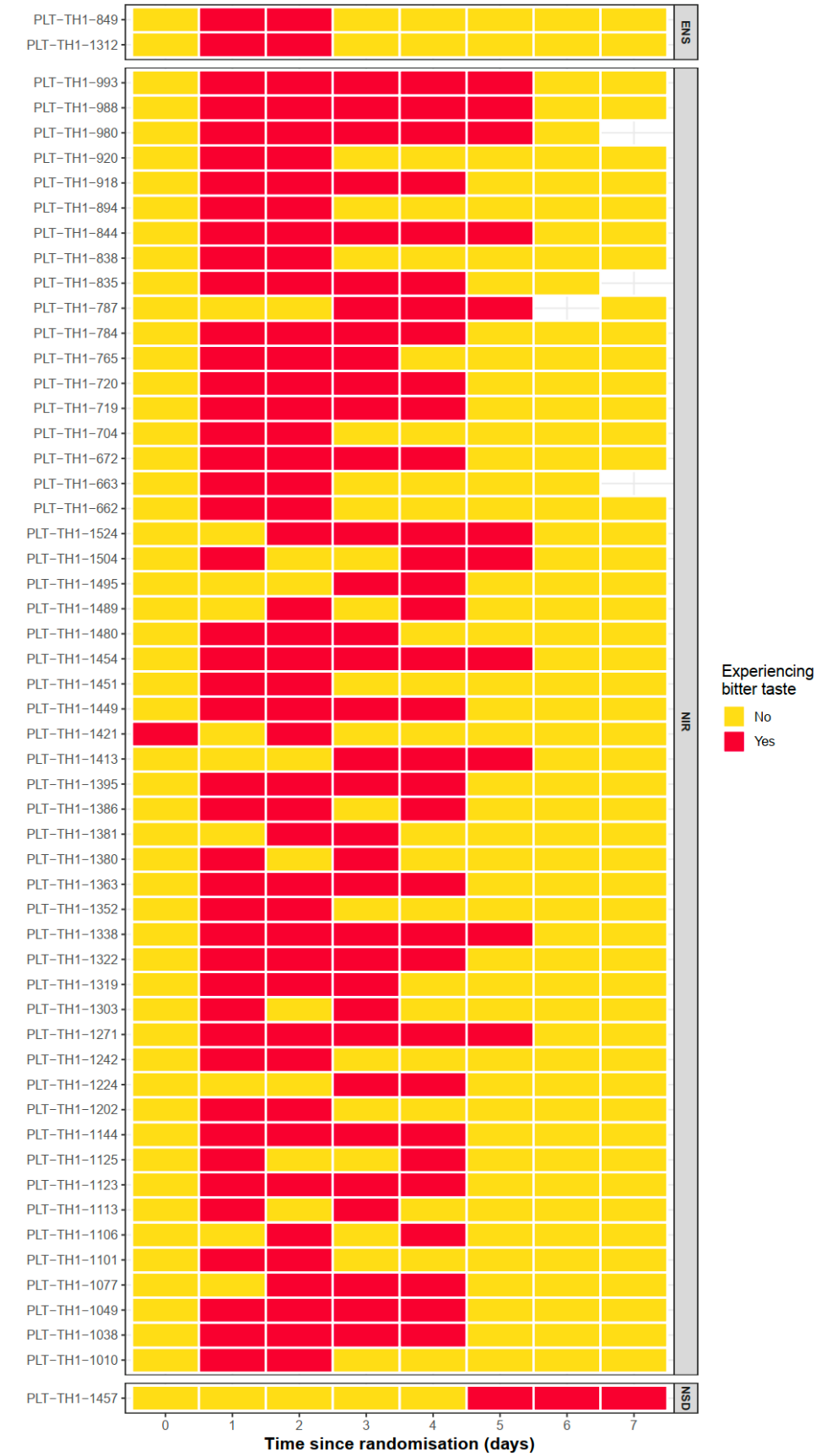

**Figure S5:** Occurrence of patients reporting dysgeusia (bitter/metallic tastes) in the enstrelvir (ENS), ritonavir-boosted nirmatrelvir (NIR), and no study drug (NSD) groups.

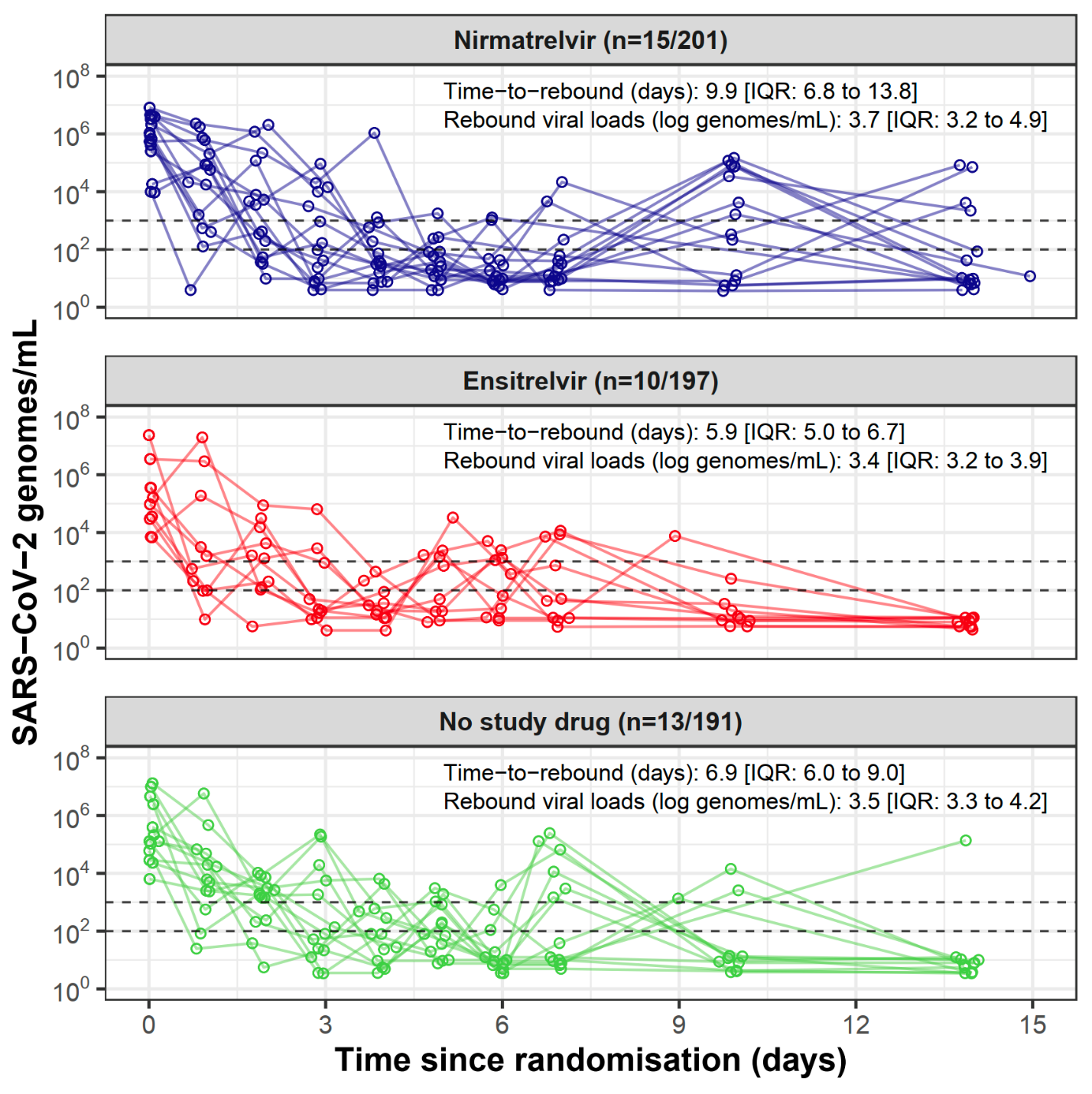

***Figure S6:*** *Viral rebound. Individual oropharyngeal viral load profiles in patients who had a viral rebound under the pre-specified definition (viral load <100 genomes/ml for > two consecutive days, followed by a viral load >1000 genomes/ml at any later timepoint).*

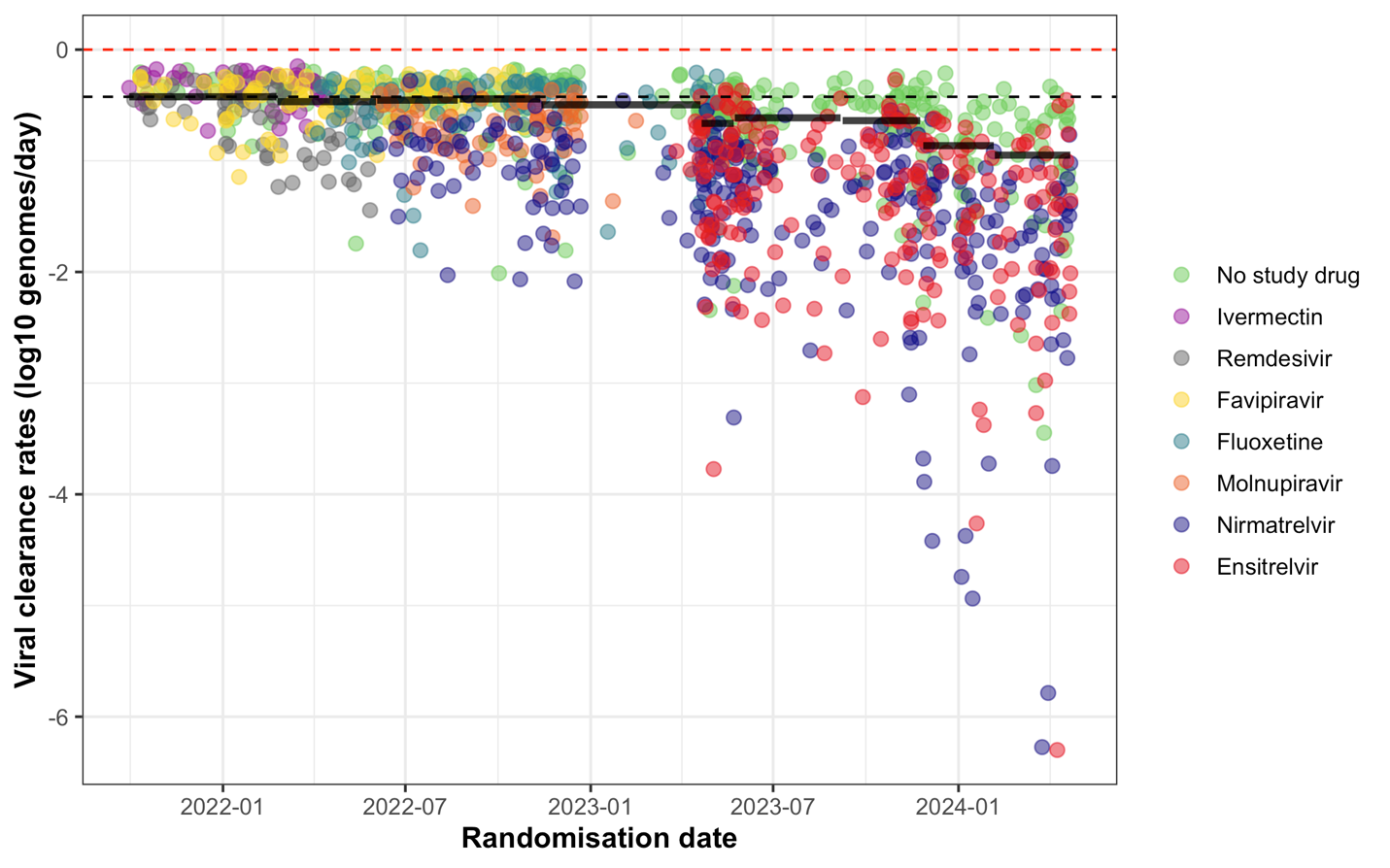
***Figure S7:*** *Estimated viral clearance rates (slope parameters) from the individual patient meta-analysis, adjusted for the effects of calendar time by treating the time period as a random effect on the slopes, showing a trend of acceleration in viral clearance over time. The time periods were classified to have approximately equal patient numbers. Baseline population viral clearance rates for each period are shown as solid horizontal lines, with the horizontal black dashed line indicating the population viral clearance rate at the beginning of the PLATCOV trial (the first time period).*
