## Supplementary material for "Antiviral efficacy of oral ensitrelvir versus oral ritonavir-boosted nirmatrelvir in COVID-19": SAP

### Finding Treatments for COVID-19: A Trial of Antiviral Pharmacodynamics in Early Symptomatic COVID-19 (PLATCOV): Statistical Analysis Plan

March 5, 2024

Registered at clinicaltrials.gov number **NCT05041907**

**Version 4.0**; refers to Master Protocol version 6.0

**Written by:**
Dr James Watson (Statistician)
Signature: Date: 5th March 2024

**Reviewed by and approved by:**

Prof Sir Nicholas White (Co-PI)
Signature: Date: 6 March 2024

Dr William Schilling (Co-PI)
Signature: Date: 5th March 2024

**S3 Casirivimab/imdevimab** 27

**S4 Tixagevimab/cilgavimab** 27

### 1 Summary of changes since version 1

The PLATCOV trial design has evolved as information has accrued and treatment policies and practices have changed in response to the evolving COVID-19 pandemic and increasing availability of specific therapies.

**Version 2.0** The first interim analysis was triggered on the 2nd of March 2022 when qPCR data from the first 50 patients was received. Following the first interim analysis, the Trial Steering Committee and the Data Safety and Monitoring Board recommended changing the minimum efficacy threshold from a 5% increase in viral clearance rate to a 12.5% increase. This change would result in a lower number of patients required to make a futility decision and a larger number of patients required to make a success decision for treatments with no antiviral effects and antiviral effects similar to those estimated from published data on molnupiravir, respectively.

We also changed the primary analysis from modelling the viral load on the CT scale with batch effects to modelling the viral load on the log copies per mL scale, with standard curve transformation done using control samples from each batch. The interim analysis suggested this was more computationally stable and easier to interpret.

**Version 2.1** We have added a section regarding the primary analysis of the pharmacokinetic data gathered from all patients.

**Version 2.2** We have added a section regarding the analysis of the time to resolution of fever, time to resolution of symptoms, and proportion with normal lymphocyte counts. We updated the current status of the trial (interventions being randomised and the current positive control).

**Version 3.0** Version 3 reflects a major update to the trial design and analysis. The following major changes were made:

- We have added a non-inferiority comparison relative to the positive control. Interventions which meet the success criteria are then entered into a non-inferiority comparison with the positive control using a pre-defined margin of -10%. Stopping occurs for inferiority (probability greater than 0.9 that the antiviral effect measured by the rate of viral clearance is less than the margin) or non-inferiority (probability greater than 0.9 that the effect is above the margin).
- We have increased the margin for futility/success for the comparison with the negative control (no study drug) from 12.5% to 20%. This results in smaller sample sizes required to show futility for interventions which have no effect. This change reflects the updated trial aims which are to characterise effective antivirals and compare them with the current 'gold-standard' antiviral treatments.
- We have changed the schedule of interim analyses (now done by arm, specified as every additional 10 patients and 10 controls recruited).

Minor changes are:

- We have removed the rapid serology test from the set of covariates used in the covariate-adjusted model. By May 2022 nearly all patients in Thailand were seropositive, in addition the Brazil site did not use these tests.
- We have updated the variant-specific subgroup analysis for the casirivimab/imdevimab monoconal antibody arm; and we have added a section on the variant-specific subgroup analysis for the tixagevimab/cilgavimab monoclonal antibidy arm.

**Version 3.1** We have added a section on the exploratory analysis of the plasma serology data collected. This pre-specifies an analysis looking at the determinants of viral clearance rates (baseline antibody titre); whether effective antivirals influence peak observed antibody titres; and the relationship between increases in antibody titres and rates of viral clearance.

**Version 4.0** Version 4 reflects a major change to the definition of the primary endpoint. Following an analysis of all data accrued during the trial up until September 2023, it became apparent that rates of viral clearance have substantially increased over time. In light of this change we have changed the primary endpoint (rate of viral clearance) to be based on the first 5 days of follow-up instead of the first 7 days of follow-up.
Minor changes are:

- We have included a section on the analysis of the nirmatrelvir/ritonavir+molnupiravir combination treatment. This is now a superiority comparison with the positive control (nirmatrelvir/ritonavir).
- We have included a section summarising all secondary outcome definitions in the trial.

### 2 Trial Overview

An effective, well tolerated, safe, affordable and generally available treatment of COVID-19 that prevented progression to severe disease would be of enormous global health benefit. There are many potential antiviral therapeutics for COVID-19, mainly consisting of repurposed small molecule drugs. At time of writing, however, there is strong evidence for clinical benefit only for the monoclonal antibodies (notably the monoclonal casirivimab/imdevimab in pre-Omicron SARS-CoV-2 variants [1]) and the small molecule drugs remdesivir [2], molnupiravir [3], and nirmatrelvir [4]. The clinical benefits of these interventions were determined from large phase 3 studies in high-risk individuals enrolled shortly after the onset of a symptomatic SARS-CoV-2 infection. The primary endpoint was most commonly either hospitalisation or death from COVID-19 (usually less than a 10% event rate). As vaccine availability has become widespread in addition to high levels of natural immunity from infection, conducting these types of clinical trials has become more and more difficult as the event rates for the primary endpoints become increasingly low (e.g. the PANORAMIC trial which has enrolled over 20,000 subjects, where the event rate for the molnupiravir comparison was 0.8% [5]; thus, a very large and expensive trial was still underpowered for its primary endpoint). This calls for improved study designs which allow for the assessment of antiviral interventions in an efficient manner (hundreds rather than tens of thousands of patients). Currently there is no consensus pharmacometric methodology to determine which antivirals should be prioritised for large phase 3 and 4 evaluation, and how to compare current candidates [6].

PLATCOV is a multi-country platform adaptive trial which provides a methodology for the quantitative assessment of antiviral effects in low-risk patients with high viral burdens and uncomplicated COVID-19. We choose patients with a low risk of progression as this justifies a no treatment arm; we target patients with high viral loads as this is a subgroup in whom antiviral effects can be detected and quantified more easily. We have already evaluated and stopped ivermectin [7], remdesivir [8], the monoclonal casirivimab/imdevimab [9], favipiravir [10], molnupiravir and nirmatrelvir/ritonavir (now the positive control in the study) [11], fluoxetine (unpublished), the monoclonal tixagevimab + cilgavimab (unpublished).

This statistical analysis plan (SAP) covers analyses for both the interim and the final reports. We developed the SAP following the Guidelines for the Content of Statistical Analysis Plans in Clinical Trials [12]. It includes pre-specified decision rules for continuing or stopping individual trial arms based on effectiveness or futility.

#### 2.1 Main research questions

Each clinical site in this platform trial will test multiple interventions simultaneously (depending on local regulations and drug availability). There are two control arms: (i) no study drug (negative control), this consists of no intervention other than antipyretics; (ii) nirmatrelvir/ritonavir (positive control), this can change in the future depending on pre-specified rules. Interventions of two types will be compared with the two control arms:

- Small molecule drugs or drug combinations: either novel or newly available or repurposed antiviral drugs (currently randomizing patients to nitazoxanide, ensitrelvir, nirmatrelvir/ combined with ritonavir+molnupiravir).
- Monoclonal antibodies: none currently in the trial.

There are distinct primary objectives for each intervention type. For the newly available antivirals and repurposed drugs, we want to characterise their antiviral activity by comparing viral clearance dynamics with no treatment. Many of these drugs are already used and recommended in some countries. Showing that they do not have clinically significant antiviral activity is as important as showing that they do. For interventions that do have a clear measurable antiviral activity, we need to determine how good they are so policy makers can make informed choices. We will then compare their activity to the current gold-standard antiviral treatment (since August 2022 this is nirmatrelvir/ritonavir). This will be a non-inferiority comparison using a pre-specified threshold. For combination therapies (currently nirmatrelvir/ritonavir+molnupiravir) we want to know whether they are superior in efficacy to either component.

For monoclonal antibodies included on the platform, we are primarily interested in tracking their performance over time with respect to new SARS-CoV-2 variants (comparison with the no treatment group).

The secondary objectives of this study are:

- To characterise the determinants of viral clearance in early symptomatic SARS-CoV-2 (e.g. estimate the contribution of age, baseline serology, virus genotype, and prior vaccination);
- To determine optimal dosing regimens for drugs shown to have considerable antiviral activity (where deemed feasible a pharmacokinetic-pharmacodynamic sub-study will be performed);
- To compare fever clearance time and time to symptom resolution with respect to no treatment for interventions that are shown to have a measurable antiviral effect.

The tertiary objective of this study is to characterise the relationship between viral clearance and the risk of subsequent hospitalisation or death by day 28. However, the event rate in the enrolled population is likely to be extremely low (thus far it has been 7 out of >1500 patients enrolled) so it is very unlikely that we will be able to demonstrate any link between viral clearance and progression to severe COVID-19.

#### 2.2 Outcome definitions

**Primary outcome**

- Rate of viral clearance: estimated from the $\log_{10}$ viral density derived from qPCR of standardised duplicate oropharyngeal swabs / saliva taken daily from baseline (day 0) to day 5.

**Secondary outcomes**

- Viral rebound. This is defined as an oropharyngeal eluate viral density estimate >1000 genomes per ml for at least 1 timepoint (average 2 swabs), after $\geq$2 consecutive days of average daily viral density estimate less than 100 genomes per ml. Rebound can occur **only** after stopping treatment for at least 24 hours or after day 4 if no drug is given or a single dose intervention is given).
- Time to resolution of fever, defined for the patients with a fever at baseline (at least one axillary temperature measurement within the first 24 hours from randomisation $\geq$ 37.5). Resolution of fever is defined as an axillary temperature $\leq$37.0C for both measurements taken over one day (at least 24 hours).
- Area under the temperature curve ($AUC_{temp}$), defined as the area under curve for the temperature measurements above the steady state temperature estimated from the day 10 and 14 values.
- Time to resolution of symptoms, defined as first day with no reported symptoms.

**Tertiary outcomes**

- Hospitalisation for clinical reasons up to day 28
- Long COVID: score on post-acute COVID-19 questionnaire at day 120 using a modified COVID-19 Yorkshire Rehabilitation Scale (C19 YRSm).

### 3 Study Methods

#### 3.1 Trial design

This is a multi-centre, multi-country, open label, randomised, controlled, adaptive platform trial of antiviral interventions in early symptomatic SARS-CoV-2. There are two distinct control arms. The negative control consists of no study drug other than antipyretic; the positive control is nirmatrelvir/ritonavir (this can change if better drugs are identified). Interventions currently included in the platform are: nitazoxanide, nirmatrelvir/ritonavir+molnupiravir, and ensitrelvir. We stopped randomisation to ivermectin (futility) in April 2022 [7]; remdesivir (success) in June 2022; casirivimab/imdevimab (no more drug) in October 2022; favipiravir (futility) in October

2022; molnupiravir (inferiority relative to nirmatrelvir) in February 2023; fluoxetine (reached 120 patients) in May 2023; Evusheld (tixagevimab/cilgavimab monoclonal antibody cocktail, success) in June 2023. Nirmatrelvir/ritonavir became the positive control in August 2022 when it met the success stopping rule with evidence of very strong antiviral activity. At each site there is equal allocation to all interventions but with a minimum of 20% of patients randomised to the negative control arm.

#### 3.2 Randomisation

Randomisation is performed via a centralised web-app designed by MORU software engineers using RShiny, hosted on a MORU server. Each study nurse responsible for randomising patients has unique login credentials provided by email from the main study statistician (James Watson). Randomisation sheets are pre-generated for each study site separately using blocks of size K (where K is equal to 3 times the number of interventions available at the site) with an additional 100/K% fuzziness (randomly interchanging one of the allocations per block to avoid the study nurse knowing exactly what the Kth allocation will be).

As of December 2022, we will do factorial randomisation for the small molecule drugs and the monoclonal antibodies (in sites where the monoclonals are available).

Randomisation sheets and randomisation event logs are stored on a secure Dropbox folder (professional version that has full version control) which is accessed directly by the RShiny app. Only the study statistician James Watson and the MORU IT manager have read/write access privileges to this Dropbox folder. For cross checking purposes, the randomisation app also records the patient age and sex. Each randomisation event is logged with the corresponding time-stamp and the identity of the nurse who performed the randomisation. Each time the set of interventions available at a given study site changes, a new randomisation sheet is generated, overwriting the previous one.

#### 3.3 Sample size projections

The sample size is adaptive (there is no fixed sample size). For each intervention the final sample size is based upon pre-specified stopping rules which use a futility/success margin $\lambda_1$ for comparisons with the no study drug arm, and a non-inferiority margin $\lambda_2$ for comparisons with the positive control arm. In addition, because there are theoretical boundary cases whereby the sample size needs to be very large in order to meet the stopping rule (when the effect is exactly equal or very close to the margin), there is a maximum sample size of 120 patients per intervention arm. An intervention arm is stopped before N=120 either (i) because it meets the futility criteria (relative to the no study drug arm, defined as Probability[effect $< \lambda_1$]>0.9); or (ii) because it both meets the success criteria (relative to the no study drug arm, defined as Probability[effect $> \lambda_1$]>0.9) and a non-inferiority criteria or an inferiority criteria (relative to the positive control arm, defined as Probability[effect $> -\lambda_2$]>0.9 or Probability[effect$<-\lambda_2$]>0.9, respectively). As of version 3 of the analysis plan, stopping for success requires both meeting the success rule relative to the no study drug arm and concluding either non-inferiority or inferiority relative to the positive control arm, with comparisons done in that order.

The average sample sizes thus depend on the values of $\lambda_1$ and $\lambda_2$. Given the model-dependent nature of the analysis, simulation is required to estimate the expected distribution of sample sizes for varying effect sizes. The following sections outline how values for $\lambda_1$ (and now $\lambda_2$) were selected and changed over the course of the study.

##### 3.3.1 Sample size: before trial commencement

Our previous simulation approach attempted to set a futility and success margin (along with a probability threshold) that would result in control of the type 1 error at approximately 10% and control of the type 2 error at approximately 20% (see study protocol for exact details). The simulations assumed that the decline in oropharyngeal viral loads was log-linear (linear on the cycle threshold (CT) scale), using measurement error estimated from an open access database of prospectively followed individuals with frequent viral load measurement [13, 6]. We calibrated plausible effect sizes using preliminary data from the casirivimab/imdevimab phase 2 studies (this suggested increases of approximately 20% in the slope of the viral clearance on the CT scale) [14].

The first interim analysis at 50 patients suggested that these simulations had underestimated inter-individual variability in viral clearance, and had underestimated plausible antiviral effect sizes. We therefore changed the futility/success threshold from 5% to 12.5%.

###### 3.3.2 Updated sample size calculations

In November 2022 we re-ran a series of simulations to re-assess sample size requirements and optimal stopping rule thresholds using the data from the first 500 patients enrolled in the PLAT-COV trial. The simulations assessed thresholds $\lambda_1 \in \{1.125, 1.15, 1.2\}$ (i.e. 12.5%, 15% or 20%) for the futility/success assessments relative to the no study drug arm; and thresholds $\lambda_2 \in \{0.905, 0.875, 0.85\}$ (i.e. -9.5%, -12.5%, or -15%) for the non-inferiority assessments relative to the positive control arm. We used the linear model fit to the first 500 patients to simulate trial data. 100 iterations were done for each permutation of $\lambda_1, \lambda_2$ and for three hypothesised effect sizes (0, 40% and 60% increase relative to no study drug), i.e. 2700 simulations in total. Preliminary simulations showed that most false positive or false negative results (stopping early for futility an intervention that has effect of 40 or 60%, or stopping early for success an intervention that has an effect of 0%) occurred for interim analyses done on 10 patients per arm. For this reason, we updated the interim analysis plan so that the first interim analysis is triggered at 20 patients per arm, with subsequent analyses for each additional 10 per arm. For the non-inferiority comparisons, they start when there are at least 40 patients per arm, and only if the success stopping rule has been met (Figure 1). Each model fit only used simulated data from the intervention and the relevant control arm (the negative control for the futility/success stopping rules, the positive control for the non-inferiority stopping rules).

**Choosing optimal $\lambda_1$** The results of the simulations for the success and futility stopping rules are shown in Supplementary Figures S1-S2. Comparable results were obtained as for the previous sample size simulations [6]. For $\lambda_1 = 1.125$ (12.5%) and an intervention that had no effect, the median sample size was 40 patients per arm with 4% false positive results and 28% of inconclusive trials at 120 patients per arm. Increasing $\lambda_1$ to 1.2 (20%) would reduce the median sample size to 30 patients per arm and result in approximately 1% false positive results and 18% of inconclusive trials at 120 patients per arm.

For an intervention with an effect of 40% (i.e similar to remdesivir in our trial [9]), the median sample size was 30 patients per arm for a threshold of 12.5% and 50 patients per arm for a threshold of 20%. In all cases, false negative results were less than 3% of simulations, and inconclusive results increased from 14% for a threshold of 12.5% to 34% for a threshold of 20%.

For an intervention with an effect of 60% (i.e similar to nirmatrelvir/ritonavir in our trial), the median sample size was 20 patients per arm, regardless of the $\lambda_1$ threshold. False negative results were less than 1% of simulations, and inconclusive results increased from 1% for a threshold of 12.5% to 9% for a threshold of 20%.

Following discussion with the trial steering committee, it was decided to increase the $\lambda_1$ threshold to 20% in order to stop poorly performing arms earlier (this reflects how the therapeutic priorities of the trial have changed as the pandemic has progressed).

**Choosing optimal $\lambda_2$** The results of the simulations for the non-inferiority stopping rules are shown in Supplementary Figures S3-S4.

In simulations which met the stopping rule for success (Probability[effect>$\lambda_1$]>0.9), stopping for non-inferiority or inferiority was then assessed for each of the three threshold values. Only in 1 simulation with an effect of 0% was the non-inferiority threshold met (<0.3% false positive result).

An effect of 40% relative to no study drug arm is very close to/on the boundaries defined by the $\lambda_2$ values, assuming that the positive control has an effect of 60% relative to the no study drug. For all three boundary values, around half the simulations met the non-inferiority criterion, 10% were inconclusive and 40% met the inferiority criterion.

For an effect of 60% relative to the no study drug arm (i.e. same as the positive control), 70% to 78% of simulations met the non-inferiority criterion for the -9.5% (i.e. most stringent) to the -15% (least stringent) thresholds, respectively. Around 6% of simulations were inconclusive at 120 patients, and 24 to 17% resulted in a false inferiority result for the -9.5% (i.e. most stringent) to the -15% (least stringent) thresholds, respectively.

Following discussion with the trial steering committee, it was decided to initially set the $\lambda_2$ threshold at -10%. This results in earlier stopping of interventions which are clearly less effective than the positive control and later stopping for interventions with an effect close to the positive control.

##### 3.4 Framework

All primary analyses and stopping decisions will be made using models including data from **contemporaneous controls only**. As this is an adaptive trial, temporal drift (due to mutations in the virus, changes in the patient characteristics, etc.) can confound comparisons with non-contemporaneous controls. However, we will also fit models to the full data set (using all controls), with additional covariate factors for the different trial epochs.

All primary comparisons for futility/success will be made with respect to the negative control arm; all primary comparisons for non-inferiority will be made with respect to the positive control arm (no "across intervention" arm comparisons in the primary analysis). In the case of a site not randomising patients to the negative control or the positive control (because of local objection or availability), that site will only provide indirect data to support the treatment estimates (under the hierarchical model structure). All decisions concerning efficacy will be based on super-superiority relative to the negative control arm ($\lambda_1$) whereby the probability of an effect is defined under the model as the posterior probability that the increase in viral clearance relative to the control arm is greater than $\lambda_1$ (20% from now on).

For interventions which show a measurable antiviral effect (i.e. meet the stopping rule for success), we will then compare their antiviral effect to the positive control arm (currently nirmatrelvir/ritonavir), using the non-inferiority threshold of -10% ($\lambda_2$).

#### 4 Statistical interim analyses and stopping rules

We plan frequent interim analyses (Figure 1). This is to allow for near-real time monitoring of accrued data in order to detect possible issues in patient recruitment, viral load swabbing, or with the PCR assays at the different sites. The stringent margin for success (greater than 20% increase) results in a very low type 1 error. In addition, meeting the success threshold triggers a subsequent non-inferiority comparison with the positive control arm and where deemed appropriate an intensive PK-PD sub-study to determine dose-response effects in order to gather additional pharmacometric data to inform dosing for that intervention. If there is strong *a priori* evidence of antiviral efficacy (e.g. molnupiravir, nirmatrelvir) additional dense blood samples for population PK-PD modelling will be obtained from enrolled patients willing to have daily blood sampling (all patients in the study have blood samples taken at baseline, day 3 and day 7 trough levels, and day 14).

##### 4.1 Frequency and timing of the interim analyses

Decision making analyses will be done by arm (only using the data from that arm and the concurrent negative or positive controls). From version 3 of the SAP, the first interim analysis for a given arm will be performed when a minimum of 20 patients randomised to that arm (and 20 concurrent negative controls) have available and analysable qPCR data (PCR data will lag behind actual recruitment, usually by approximately 3 weeks). Subsequent analyses will be performed for every additional 10 patients (10 randomised to the intervention and 10 to the control arm).

##### 4.2 Interim analysis decision making

Figure 1 shows the decision algorithm for removing an intervention from the platform or replacing the positive control (current nirmatrelvir/ritonavir). Only the study statistician (James Watson) will be unblinded to all interim analyses. Each interim analysis report will be sent to the Data Safety and Monitoring Board (DSMB) along with a summary of whether the futility or success thresholds for any of the interventions have been met. The DSMB will then make recommendations for continuing or stopping the intervention arms. Concurrent data from other trials (e.g. safety data) may be used in making these decisions. If an arm is stopped for futility or an intensive PK-PD study is triggered, then all the data pertaining to the negative control arm and the intervention arm

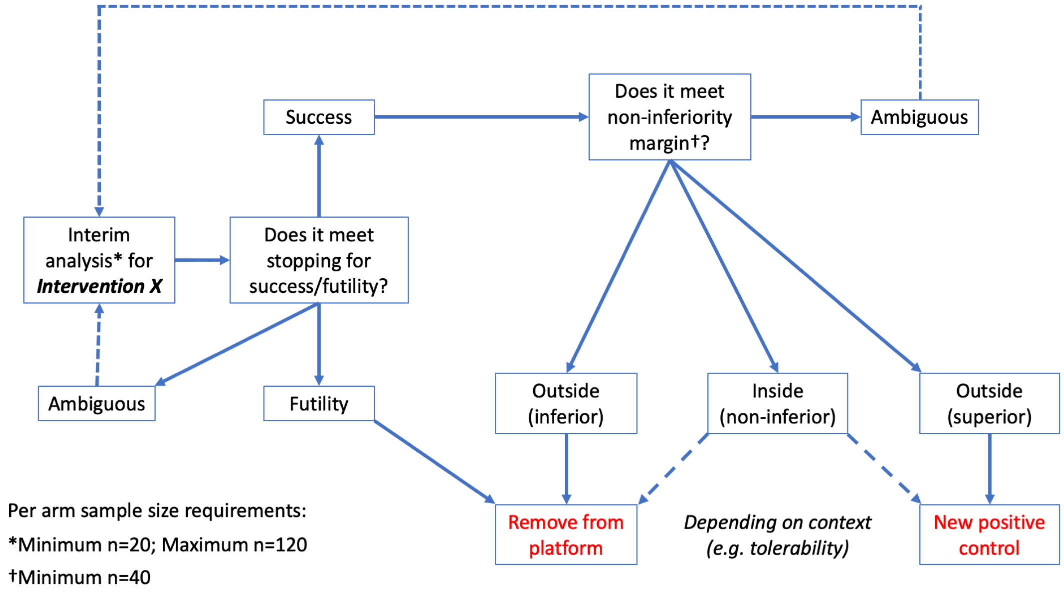

Figure 1: Planned interim analyses and decision rules for removing an arm from the platform or making an arm the new positive control.

will be presented to the Trial Steering Committee (TSC), along with the DSMB recommendations. The interim analysis of the first 50 patients with available qPCR data was presented unblinded to the TSC in order to determine necessary changes to the statistical analysis plan.

##### 4.3 Timing of final analysis

As this is a platform trial, no overall final analysis is planned. For each intervention studied, we define the "final analysis" as the point when the last patient randomized to that arm passes their day 28 follow-up.

##### 4.4 Timing of outcome assessments

For the primary and secondary endpoints we will use all viral load measurements taken up until day 5. Time will be defined as time since randomisation (units of days, including all timepoints $<$ 5.5 days since randomisation as there will be some variation in exact clock times of the follow-up swabs). This is a change from previous versions where we use data up until day 7, see Appendix S2 for rationale.

For rebound, this can occur only after day 4 (i.e. day 5 or later) for the no study drug arm, and at least 24 hours after stopping drug for the treatment arms.

For fever and symptom resolution, time to resolution will be calculated from daily measurements taken over the first week with right censoring for unobserved events (withdrawal or no resolution by 7 days).

The tertiary endpoint all cause hospitalisation will be taken up until day 28. The tertiary endpoint for long COVID will be taken at approximately day 120 (+/- 1 month).

#### 5 Statistical Principles

##### 5.1 Posterior estimates

Treatment effects will be estimated under a Bayesian framework using a hierarchical model with weakly informative prior distributions. For the hyperparameters, the prior distributions are chosen for computational reasons to aid model convergence. For the key population level parameters, we will use priors based on analysis of open access data [6]. The prior on the treatment effect is driven by plausible maximum effect sizes given published data on viral decreases in patients randomised to casirivimab/imdevimab or placebo [1].

##### 5.2 Adherence and protocol deviations

This study is open label and thus it is possible that some patients who are randomized to the no study drug arm, or to study interventions perceived to be ineffective, may be given alternative rescue treatment if their symptoms persist and the treating physician is worried that they will deteriorate clinically. This can introduce confounding between the treatment allocation and outcome. Notably, if multiple patients randomized to the negative control were in fact given the positive control before day 7, this could impact the viral loads during follow-up and thus bias the estimates of the rate of viral clearance in the negative control arm. For all protocol deviations regarding treatment (either stopping treatment for safety reasons or switching treatment arms), we will consider all viral load measurements taken after the deviation as censored, i.e. we will estimate viral load clearance rates only using the data during the period of protocol adherence. We will summarise the number of treatment protocol deviations by site.

##### 5.3 Analysis populations

The primary analysis will be in a modified intention-to-treat (mITT) population, including follow-up data only from the period of treatment adherence. A minimum of 3 days of follow-up data are necessary in order to be included in the mITT population (e.g. patients who discontinue before day 2 will not be included).

An optional secondary analysis can be performed in the per-protocol population, defined as patients who had 100% adherence to the allocated intervention arm. Specific reasons for treatment discontinuation will be outlined and recorded in order to assess confounding bias. This will only be done in specific cases where there is a good reason for discontinuation unrelated to antiviral effect (i.e. side-effect such as gastrointestinal issues as seen with high dose nitazoxanide).

The safety population will include all patients who have received at least one dose of the intervention.

#### 6 Trial population

Figure 2 shows an example CONSORT diagram summarising the number of patients screened; the reasons for exclusion; the number of enrolled patients randomized to each arm; and the number of patients who complete treatment and follow-up and who are included in the modified intention to treat analysis (mITT).

##### 6.1 Screening data

We will summarise the reasons for exclusion for the screened patients not eligible for enrolment. This will not be done for the interim analysis, only for the final analysis once an arm is stopped for futility or success.

##### 6.2 Eligibility

The key trial eligibility criteria are as follows:

- Previously healthy adults, aged 18 to 60 years (low risk of developing severe COVID-19);
- SARS-CoV-2 positive by lateral flow antigen test within 2 minutes (suggesting a high viral load) OR a positive PCR test for SARS-CoV-2 within the last 24 hours with a CT value of less than 25 (all viral targets);
- Symptoms of COVID-19 (including fever, or history of fever) for less than 4 days (96 hours);
- Oxygen saturation $\geq$96% measured by pulse-oximetry at time of screening.

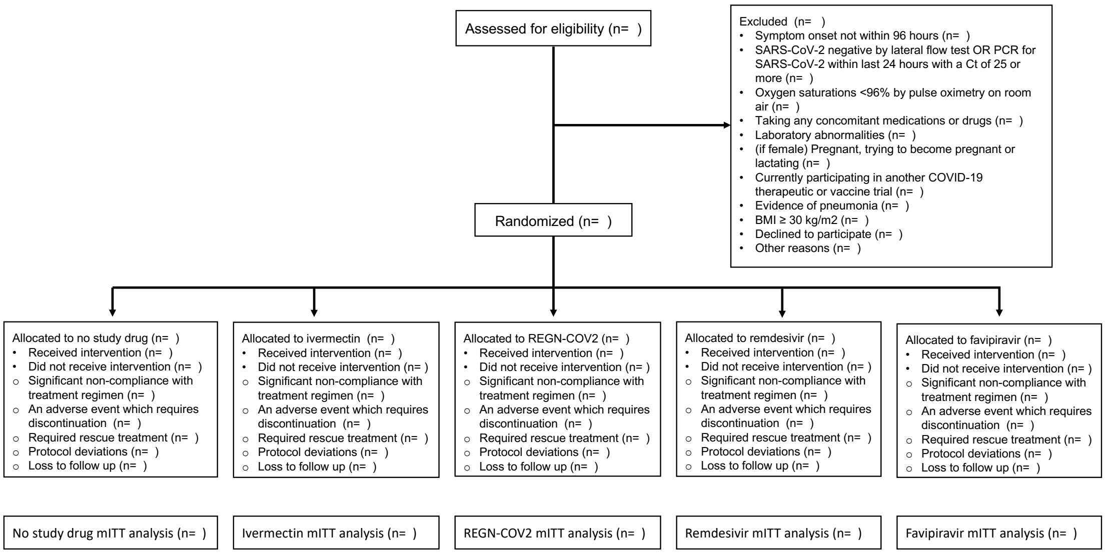

Figure 2: Example CONSORT diagram summarising screening, reasons for excluding patients and randomisation.

#### 6.3 Withdrawal and follow-up

Level of withdrawal will be tabulated and cover the following aspects:

- Discontinuation from any of the study interventions
- Withdrawal from study follow-up
- Withdrawal from entire study and requests that data are not used.

Data will be tabulated for the timing of withdrawal from follow up or lost to follow up. The number of withdrawals and reason will be recorded. A participant may withdraw from the intervention, or be withdrawn for the following reason.

- Withdrawal of consent by participant
- Alteration of participants circumstances or condition which gives justification for discontinuation as decided by investigator.

A table will summarise the loss to follow up and withdrawals during the study with the corresponding reasons.

#### 6.4 Baseline patient characteristics

The following key baseline characteristics will be summarised, stratified by site:

- Age and sex;
- Baseline oropharyngeal viral load (in copies per mL: the mean value of the 4 swabs taken at randomisation);
- Baseline quantitative serum antibody titres (when available);
- Vaccination status (vaccine type, number of doses: expressed as not vaccinated, partially vaccinated and fully vaccinated);
- Days since onset of symptoms.

These will be summarised by their mean or median and standard deviation/range for the continuous variables and by their mean for the binary variables.

### 7 Analysis

#### 7.1 Outcome definitions

The primary outcome is the inferred rate of oropharyngeal viral clearance expressed as a slope coefficient (the mean posterior estimate and the 95% credible interval) for the daily change in the log viral load (on the $\log_{10}$ copies per mL scale). This will be inferred under a Bayesian mixed effects (hierarchical) model using the $\log_{10}$ viral load measurements up until day 5. The observed CT values will be transformed to viral genome copies per mL using the set of control samples from each batch.

##### 7.1.1 Viral load quantification

All viral load quantification will be done using ThermoFisher TaqCheck™ SARS-CoV-2 Fast PCR assay. Each qPCR assay is run on a 96 well plate, composed of all the aliquots from the transport media of the swabs from 4 patients (10 timepoints each done in duplicate, i.e. 20 samples per patient) and one control set composed of 6 spiked control samples of known viral densities over the range $10^7$ to $10^2$ viral copies per mL, each done in duplicate (12 in total). For each plate (i.e. batch) we can thus estimate a standard curve (which defines the conversion from the CT value to a number of viral copies per mL). At each interim analysis we will perform a quality control check by comparing the estimated standard curves from each plate, both within sites and across sites (each country will run the qPCR assays separately). In addition, we will use the standard curve data to estimate heteroscedasticity in the viral load estimation. We expect the qPCR measurement error to increase as the viral load decreases. Heteroscedasticity can be shown visually by plotting the true viral load densities in the control duplicated samples against their observed difference in CT value. We can then estimate the variance in the differences as a function of the true viral density.

To help adjust for variation in the human cell content of the swabbed sample the qPCR method assesses viral densities and also human cell densities using RNaseP as a human cell marker.

#### 7.2 Analysis Methods

##### 7.2.1 Statistical model for the analysis of the serial viral load data

The primary analysis will consist of fitting Bayesian hierarchical (mixed effects) linear models to the $\log_{10}$ oropharyngeal viral load data up until day 5. The treatment effect is defined as the proportional change (expressed as a multiplicative term) in the population slope of the daily change in log viral load. We will model the data on the copies per mL scale, after conversion from CT values using controls from each 96 well plate. The standard curve transformation will be done by fitting a linear mixed effects (random slope and random intercept for each plate) model to the control data: regressing the CT values on the known log viral densities. This borrows information across plates but allows for batch effects.

The general model likelihood takes the following form:

$$y_{i,t,k} \sim \text{Student}\left(\lambda, \alpha_0 + \alpha_i + \alpha_k + \alpha_{cov} + \gamma x_{i,t} + \beta_0 e^{\beta_k + \beta_i + \beta_{cov} + \beta_{T(i)}} t, \sigma^2\right)$$

where:

- $y_{i,t,k}$ is the log viral load ($\log_{10}$ copies per mL) for patient $i$ at time $t$ (in days since randomisation) from site $k$.
- $T(i)$ is the randomized treatment allocation for individual $i$.
- $\sigma^2$ is the variance of the measurement error.
- $\lambda$ is the degrees of freedom of the student-t error model.
- The slope of viral clearance decomposes into 5 multiplicative terms: the population mean slope $\beta_0$; the site random effect $\beta_k$ ($e^{\beta_k}$ is thus the proportional change in slope); the individual random effect $\beta_i$; the sum of the covariate effects $\beta_{cov}$; and the treatment effect (fixed across sites and individuals) $\beta_{T(i)}$. All comparisons are made relative to the no antiviral treatment control arm, so we set $\beta_{control} = 0$.

- The intercept term (baseline log viral load) decomposes into 4 terms: the population intercept $\alpha_0$; the site random effect $\alpha_k$; the individual random effect $\alpha_i$; and the sum of any covariate effects $\alpha_{cov}$.

- $x_{i,t}$ is the relative human RNaseP quantification (RNaseP ΔCT value: this is proportional to the log number of human cells) for patient $i$ at time $t$. The parameter $\gamma$ thus provides an adjustment for human cell content in each swab. We scale $x_{i,t}$ so that it has mean 0.

The analysis will use three model types. The primary model is the linear model with adjustment for virus variant and site. A secondary model then incorporates the covariate adjustment terms $\alpha_{cov}$ and $\beta_{cov}$ (see below for their definition). If analysis only has data from one site, then the site random effect term $\alpha_k$ will be dropped. The final model (model 3) is a non-linear version of models 1 and 2 (see below). Efficacy estimation is made based on model 2 (log-linear model with covariate adjustment).

We have chosen a student-$t$ distribution for the model likelihood (with the number of degrees of freedom $\lambda$ estimated from the data) as this is robust against departures from normal (Gaussian) error, and against model mis-specification, notably concerning the assumption of log-linear decline in viral loads. Prior analyses carried out in order to prepare this statistical analysis plan suggest that a considerable number (e.g. up to 5%) of viral load densities can depart substantially from the expected distribution under a simple log-linear model with Gaussian error [6]. This can be explained partially by variation in the recovery from oropharyngeal sampling and partially by the patterns of bi-exponential decay in viral loads. It is unclear if the slope of the second phase of elimination in a bi-exponential decline is affected by interventions. We therefore choose not to fit bi-exponential models as this makes the interpretation of the treatment effect more difficult and prior modelling suggested no major impact in terms of treatment effect estimation [6].

Goodness of fits will be assessed using leave-one-out validation [15] and Bayesian $R^2$ approximation [16].

###### 7.2.2 Covariate adjustment

In addition to the human RNaseP adjustment, the primary analyses will adjust for the following covariates (through the parameters $\alpha_{cov}, \beta_{cov}$):

- Age (in years - scaled to have mean zero);

- A quantitative measure of serological status using a pre-specified antibody or antibodies (not yet decided). These data are not yet available.

- Previous SARS-CoV-2 vaccination (as an ordinal variable: 0: no doses; 1: partially vaccinated with only one dose: 2: fully vaccinated with 2 doses; 3: 3 or more doses);

- The SARS-CoV-2 lineage or sub-lineage. The reference strain is the Delta lineage, with Omicron sub-divided into descendant lineages (BA.1, BA.2 etc). Given the difficulty in pre-specifying exact subgroupings, we allow some flexibility (e.g. considering BA.2.75 as separate from previous BA.2 lineages).

- The trial "epoch" (only for analyses including non fully overlapping intervention arms): this is defined as the discrete intervals of time at which new interventions enter or exit the study. This is to adjust for temporal effects which could confound across arm comparisons made for non-contemporaneously recruiting arms.

If any of the binary covariates show no variation (e.g. all patients are vaccinated) then we will drop the term from the model. We will fit both the covariate adjusted model and the non-adjusted model in all instances as to check for sensitivity to the covariate model. A sensitivity analysis regarding temporal drift will be done by analysing each arm individually along with contemporaneously enrolled controls.

###### 7.2.3 Resolution of fever and symptoms

Time to resolution of fever and time to resolution of symptoms will be analysed by comparing Kaplan-Meier curves (right censored data). Patients are defined as having a fever at baseline if at least one axillary temperature measurement within the first 24 hours from randomisation is

$\geq 37.5$. Switching to aural temperature measurement is under consideration in order to reduce measurement variation. Resolution of fever is defined as an axillary temperature $\leq$37.0C for both measurements taken over one day (at least 24 hours). Resolution of symptoms is defined as no reported symptoms. An additional analysis will look at the proportion of patients with lymphocyte count within the normal ranges at D3 and D7. Differences in time to resolution for both endpoints will be tested using the log-rank test. Differences will be quantified by the change in the median time to resolution.

#### 7.3 Pharmacokinetic analyses

All patients will have 4 plasma samples taken at: randomisation, day 3, day 7 (all three are pre-dose), and finally day 14. Drug concentrations will be quantitated from these samples in order to explore dose-response relationships between drug (or metabolite) exposure and outcome (slope of viral clearance). There are two objectives for the pharmacokinetic data:

- First, to verify that plasma concentrations are within the expected range (using published pharmacokinetic data from equivalent patient groups and dosing);
- Second, to explore whether there is any correlation between plasma concentrations and outcome (rate of viral clearance).

The first objective will use published pharmacokinetic data to assess whether drug levels are within the expected range. For the second objective, the primary analysis will be performed using the day 3 level (a trough level). We will fit a linear model between the day 3 drug (and/or active metabolite) level (log transformed) and the mean viral clearance slope estimate from the main analysis in patients allocated to the intervention arm. Significance is defined as a two-sided test at the 5% level. If appropriate more complex population pharmacokinetic modelling will be performed to reconstruct individual patient concentration profiles in order to relate to the viral clearance rate.

#### 7.4 Subgroup analyses

The primary pre-specified subgroup analysis will examine the relationship between the efficacy of the monoclonal antibodies (previously the Regeneron antibody cocktail casirivimab/imdevimab [9], currently tixagevimab/cilgavimab) and the virus genotype. Previously we had pre-specified subgroups based on the major WHO lineages, however, given the recent diverging trends in SARS-CoV-2 evolution, this approach does not make much sense anymore. Instead we will use *in vitro* assay data to define a set of canonical non-synonymous amino-acid changes which are predicted to change substantially the efficacy of the monoclonal in question. For each monoclonal in the platform, we will write a specific appendix outlining exact subgroup definitions (amino acid calls are based on the nanopore sequencing data).

Subgroup analyses will be done by fitting an interaction term $\delta x_{var}(i)e^{\beta_{T(i)}}$ for the relevant $T(i)$; where $x_{var}(i)$ is the virus genotype defined subgroup infecting individual $i$.

Secondary subgroup analyses will be carried out only for treatment arms that show an antiviral effect (meet the success criteria). We will look at subgroup effects for (listed in order of priority):

- SARS-CoV-2 variant of concern;
- Serological status on admission (using quantitative serum antibody or antibodies);
- Vaccination status;
- Age and sex;

For the analysis of tixagevimab/cilgavimab, see Supplementary section S4.

#### 7.5 Characterising determinants of viral clearance: serology

Stored plasma samples are taken on day 0, and then on days 3, 7, and 14. Assays will include a standard ELISA assay for the ancestral SARS-CoV-2 strain (quantitative S-protein IgG), and neutralising antibody tests using later variants matched to the variant/subvariant the patient was infected with. These antibody (Ab) measurements (titres) (S-protein IgG or neutralising antibody titre or other antibodies) will be used to answer the following questions:

1. What is the relationship between baseline Ab titre(s) and rate of viral clearance?
2. Does baseline Ab titre correlate with peak viral load?
3. Is there a relationship between baseline Ab titre and clearance of virus in patients randomised to the monoclonal antibody therapy arms?
4. Do effective antivirals (eg nirmatrelvir) attenuate the antibody response assessed at D14 ?
5. What is the relationship between the change in Ab titres (between days 0 and 7) and the rate of viral clearance?
6. What is the relationship between day 7 Ab titre, drug treatment and viral rebound?

Questions 1-3 involve adding baseline Ab titres as explanatory covariates for the slope (ie rate of clearance) and intercept (ie baseline viral load) in the main analysis models. Question 4 will be addressed sing a simple linear regression onto the day 14 Ab titres, with baseline Ab titres and randomised treatment as the explanatory covariates. Question 5 will be addressed by comparing estimated rates of clearance under the model with estimated changes in Ab titres (correlation). Question 6 considers whether day 7 serology predicts viral rebound (especially of interest for ritonavir-boosted nirmatrelvir).

We use a pragmatic (non-model based) definition of rebound: daily viral load estimate (median of two daily samples) less than 100 genomes per ml for at least two consecutive timepoints, followed by a daily viral load estimate greater than 1000 genomes per ml for at least 1 timepoint after. This definition is similar to that used in [17].

#### 7.6 Prior distributions

In all the following, for the normal distributions, the second term corresponds to the scale (standard deviation not the variance). The viral load values are on the $\log_{10}$ scale (i.e. for the intercept term 6 would correspond to 1,000,000 copies per ml).

**Population level parameters**

$$\alpha_0 \sim \text{Normal}(6, 2) \quad \text{(population intercept)}$$

$$\beta_0 \sim \text{Normal}(-0.5, 1) \quad \text{(population slope)}$$

$$\sigma \sim \text{Normal}(1.5, 3) \quad \text{(standard deviation of measurement error)}$$

$$\gamma \sim \text{Normal}(0, 1) \quad \text{(human RNaseP adjustment)}$$

$$\beta_{T(i)} \sim \text{Normal}(0, 0.5) \quad \text{log treatment effect (subgroups also)}$$

**Covariate effects**

$$\beta_{cov} \sim \text{Normal}(0, 1) \quad \text{(covariate effect on slope)}$$

$$\alpha_{cov} \sim \text{Normal}(0, 1) \quad \text{(covariate effect on intercept)}$$

**Random effects and hyperparameters**

$$\Omega \sim \text{Cholesky}(2) \quad \text{(correlation matrix for individual random effects)}$$

$$\Sigma \sim \text{Exponential}(1) \quad \text{(standard deviation for individual random effects)}$$

$$\lambda \sim \text{Exponential}(1) \quad \text{(t-distribution degrees of freedom)}$$

#### 7.7 Sensitivity analyses

Sensitivity to the prior specification will be assessed by multiplying all prior standard deviation terms by 5. Sensitivity to the assumption of log-linear decline will be assessed by fitting an 'up-down' model of viral dynamics [18] (where "up" refers to viral multiplication and "down" the subsequent clearance). Under this model the likelihood is (dropping batch, site, and covariate terms for notational simplicity, these are added analogous to the previous log-linear model) given by:

$$y_{i,t} \sim \text{Student}\left(\lambda, \alpha_0 + \alpha_i + \log\left\{\frac{\beta_1^i + \beta_2^i}{\beta_2^i e^{-\beta_1^i[t-t^i_{\text{max}}]} + \beta_1^i e^{\beta_2^i[t-t^i_{\text{max}}]}}\right\}, \sigma^2\right)$$

Under this model, the term $\alpha_0 + \alpha_i$ is the individual peak log viral load which occurs at time $t^i_{\text{max}}$ (this is the time relative to randomisation, if negative it is therefore unobserved). The parameters $\beta_1^1, \beta_2^i$ are the growth and clearance rates for individual $i$, respectively.

If some patients are enrolled very early in the course of their illness, their viral loads may increase over the first days of follow-up. This would mean that the clearance rate estimated under a simple log-linear decline model would be biased towards a shallower slope. Randomisation protects against confounding bias as the sample size increases, however for small sample sizes unequal allocation of 'pre-peak' versus 'post-peak' individuals could influence treatment effect estimates. Treatment effects are inferred in the same way as for the log-linear model (proportional change in the population slope coefficients), by acting on $\beta_2$ (note that in reality, the treatment effect most likely would change all three parameters $\alpha_0, \beta_1, \beta_2$, however this would mean inferring three separate treatment effects as the model is non-mechanistic).

#### 7.8 Missing data

Missing viral load data will not be imputed. The model will be fit to all available viral load densities up until day 5 or day of treatment deviation. For all analyses, missing data of the key baseline variables (vaccination, virus genotype, antibody rapid test) will be imputed at random using the observed site-specific covariate distribution multiple times (5 times, the model will be refit). We do not expect much missing data (eg $<1\%$). If delays in genotyping result in missing variant data, we will impute the variant data using genetic surveillance data.

#### 7.9 Harms

Safety analyses will include all patients who have received at least one dose of the intervention. The safety and tolerability data will be pooled from all the sites that receive the same study intervention. The safety and tolerability will be assessed by comparing the frequency of adverse events and serious adverse events when compared to the control intervention.

All adverse event summaries will refer to treatment emergent adverse events, i.e. adverse events that newly started or increased in intensity after study drug administration (or from hour 0 in the control group receiving no intervention). Adverse events will be graded according to CTCAE V5.0. Adverse Events (AEs) of grade 3 and above will be recorded, the grading is as follows - 1 = mild, 2 = moderate, 3 = severe, 4 = life-threatening, 5 = fatal. All Serious Adverse Events (SAEs) will be recorded, a serious adverse event is any untoward medical occurrence that results in death, is life threatening, requires inpatient hospitalization or prolongation of existing hospitalization, results in persistent or significant disability/incapacity or consists of a congenital anomaly or birth defect.

We will tabulate AEs of grade 3 and higher, SAEs, and deaths, by treatment arm with comparisons made relative to the no study drug arm.

None of the drugs evaluated will be pre-registration or pre-approval, so adverse effect profiles for each are already well known.

#### 7.10 Statistical software and analysis implementation

All analyses will be be in done in R. Interim analyses and the final analyses will be done using RMarkdown scripts, pre-coded to ensure full transparency. All code for the statistical analysis is available on the following github repository: `https://github.com/jwatowatson/PLATCOV-SAP`. This ensures full version control by tracking all changes made to the analysis code.

The statistical models for the analysis of the serial viral load data are written in *stan* and fitted via the *rstan* interface [19]. These are available on the github repository. For each model, we will run 6 parallel chains for 10,000 iterations, discarding half for burn-in and thinning every 40 (thus 1,000 posterior draws).

Convergence of Bayesian fits will be assessed visually by examining the traceplots of the key model parameters and assessing the Rhat values for each parameter (Gelman-Rubin statistic).

### S1 Updated sample size simulations

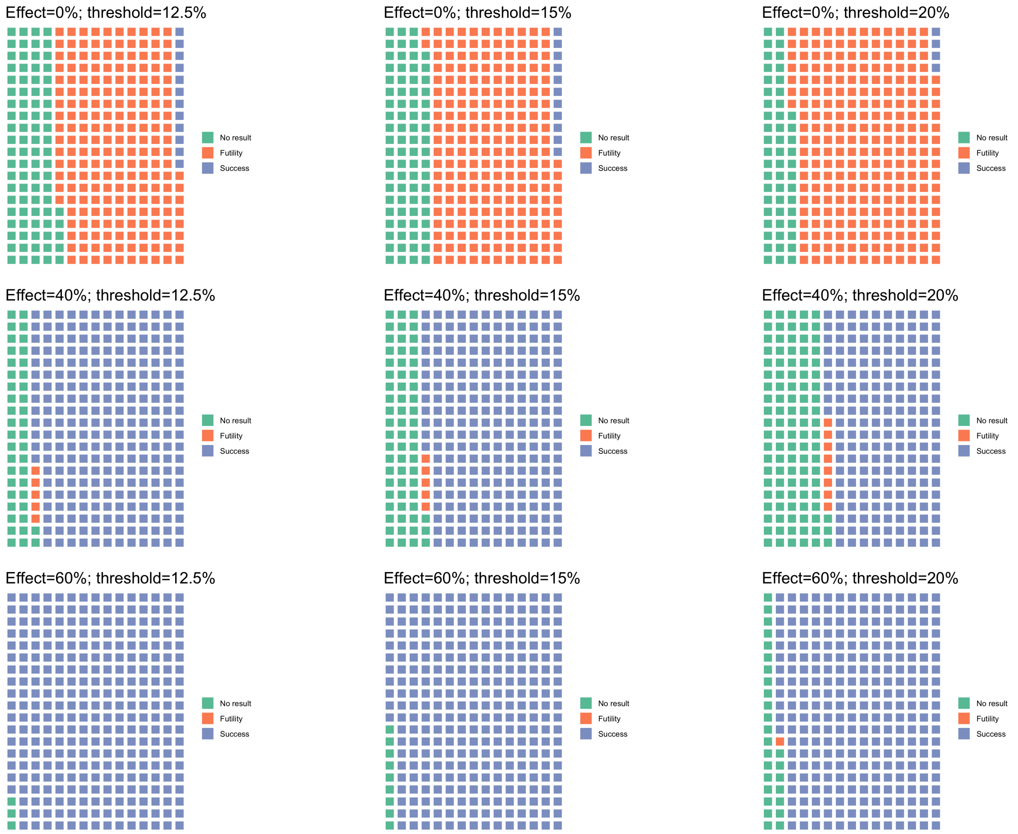

Figure S1: Waffle plots showing the proportion of each outcome (success: blue; futility: orange; no result by 120 patients: green) for the success versus futility stopping rule. 100 simulations were run for each permutation of the effect size and $\lambda_1$ threshold.

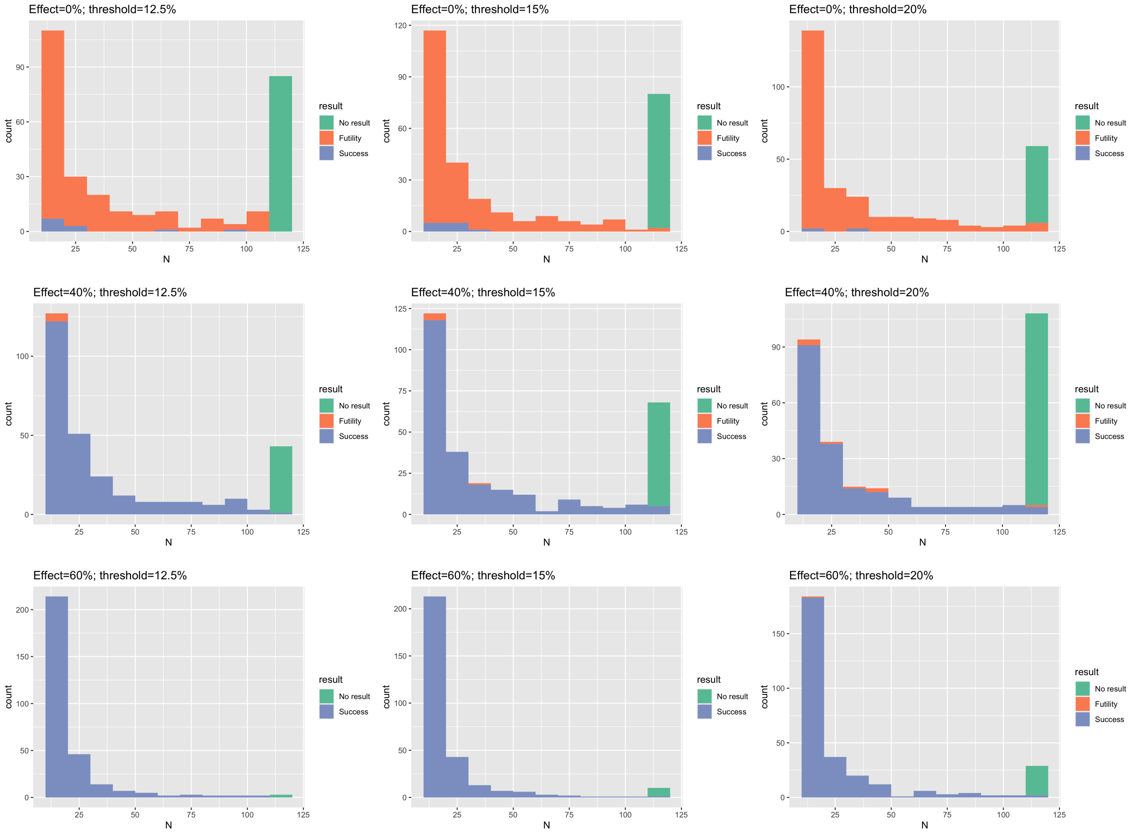

Figure S2: Histogram of sample size until stopping rule is met, same colors as in Figure S1.

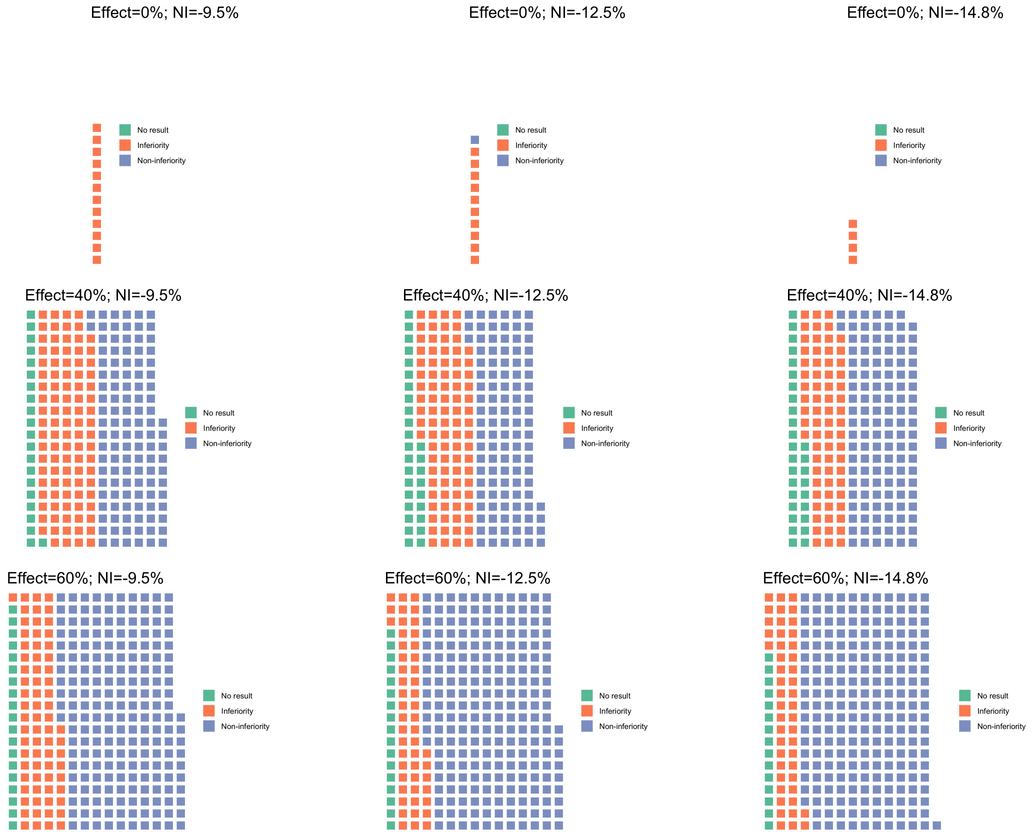

Figure S3: Waffle plots showing the proportion of each outcome (non-inferiority: blue; inferiority: orange; no result by 120 patients: green) for the non-inferiority stopping rule. 100 simulations were run for each permutation of the effect size and $\lambda_2$ threshold.

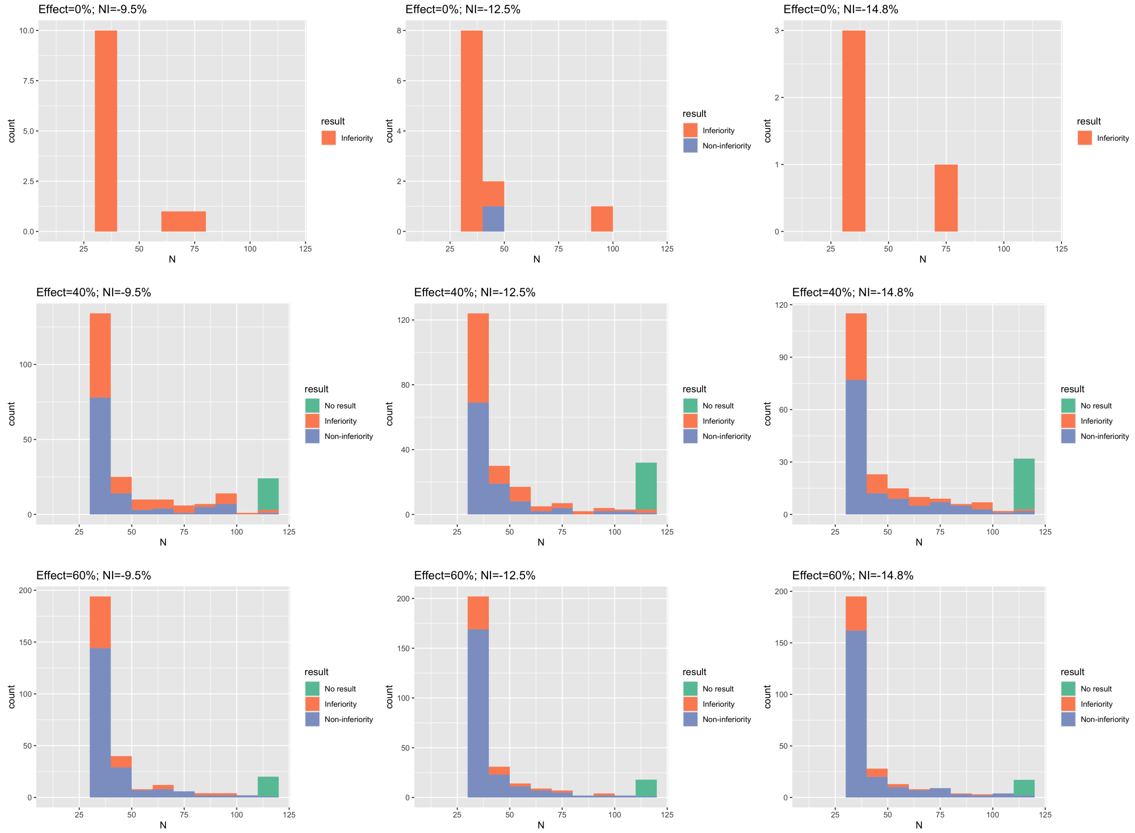

Figure S4: Histogram of sample size until stopping rule is met, same colours as in Figure S3.

### S2 Optimal follow-up duration for estimation of the clearance rate

#### S2.1 Background

In versions 3 and under of the statistical analysis plan, the rate of viral clearance was estimated using data up until day 7 under a log-linear decay model. The rationale for only using data up until day 7 was that this focuses on the initial decay phase of the bi-phasic viral clearance [6]. In October 2023 we observed that SARS-CoV-2 viral clearance rates had been steadily increasing in the no study drug arm over time. The clearance half-life had gone from $\sim$ 16.8 hours in September 2021 to $\sim$ 9.3 hours in October 2023 (Figure S5). Therefore, the clearance profiles in recent recruitment may enter their second phase (slow phase) of the bi-phasic decay quicker than their early recruitment counterparts. As a result, using a follow-up duration of 7 days, which was designed based on early viral dynamics profiles, for individuals in recent recruitment may result in reduced statistical power, given the log-linear decay model. Therefore, we explored here the optimal follow-up duration that maximises statistical power in estimating treatment effects using the log-linear decay model.

#### S2.2 Methods

We explored the follow-up duration that maximises statistical power using a bootstrap approach. We look at 6 contrasts, for treatment arms which have previously demonstrated clear antiviral efficacy (i.e. we know that they accelerate viral clearance relative to the no study drug arm):

- Remdesivir vs No study drug
- Molnupiravir vs No study drug
- Casirivimab/imdevimab vs No study drug
- Nirmatrelvir/Ritonavir vs Molnupiravir
- Nirmatrelvir/Ritonavir (before Feb 2023) vs No study drug
- Nirmatrelvir/Ritonavir (after Feb 2023) vs No study drug

The complete viral dynamics dataset was filtered to include subsets with various follow-up durations (2, 3, 4, 5, 6, 7, and 14 days) to estimate treatment effects. For instance, the 3-day follow-up subset contains viral load data for day 0, day 1, day 2, and day 3. In order to explore the uncertainties of estimated treatment effects, we conducted the analysis on 50 bootstrapping datasets (sampling with replacement) from these subsets. Finally, z-scores of the estimated treatment effects (effect size/standard error) for different follow-up duration in each comparison contrast were used to illustrate the statistical power. The follow-up duration giving the highest z-score maximise the power of estimating treatment effects.

#### S2.3 Results

The follow-up duration of 4 to 5 days was shown to maximise z-scores and, therefore, statistical power in estimating antiviral activities across the 5 included comparison contrasts (Figure S6).

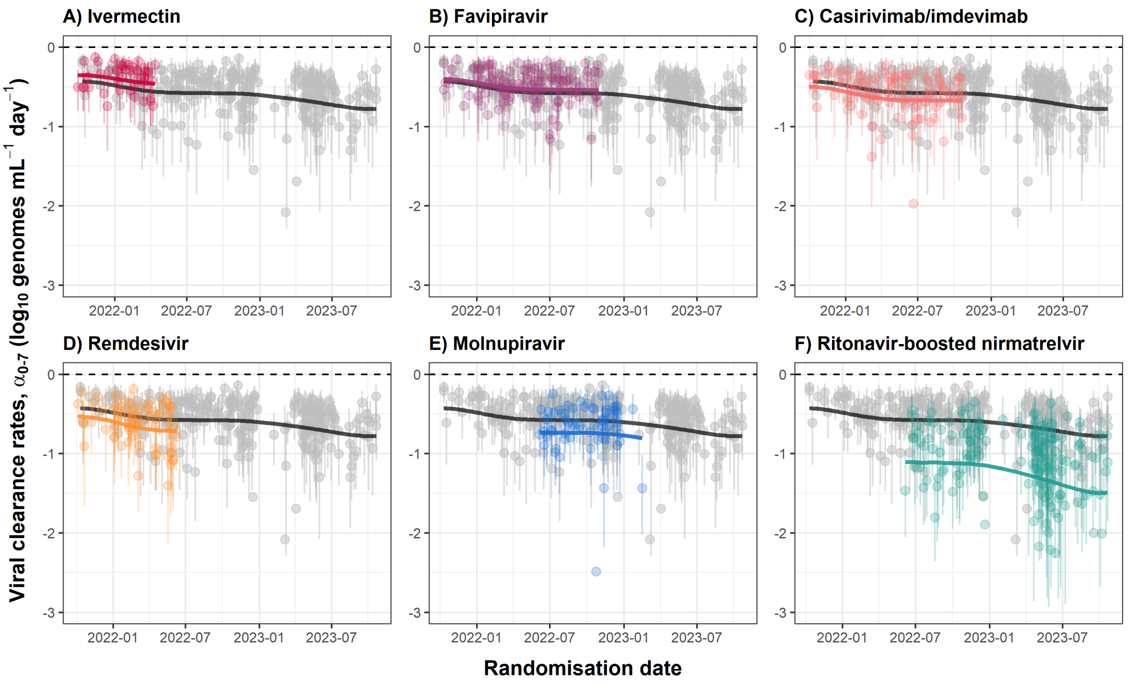

Figure S5: Individual patient rates of viral clearance between days 0 and 7. Average clearance rates for each intervention (coloured lines) and the no study drug arm (black line) are estimated from a spline fit (treatment effects parameterised as proportional change in rate). Vertical lines show 95% credible intervals under the linear model.

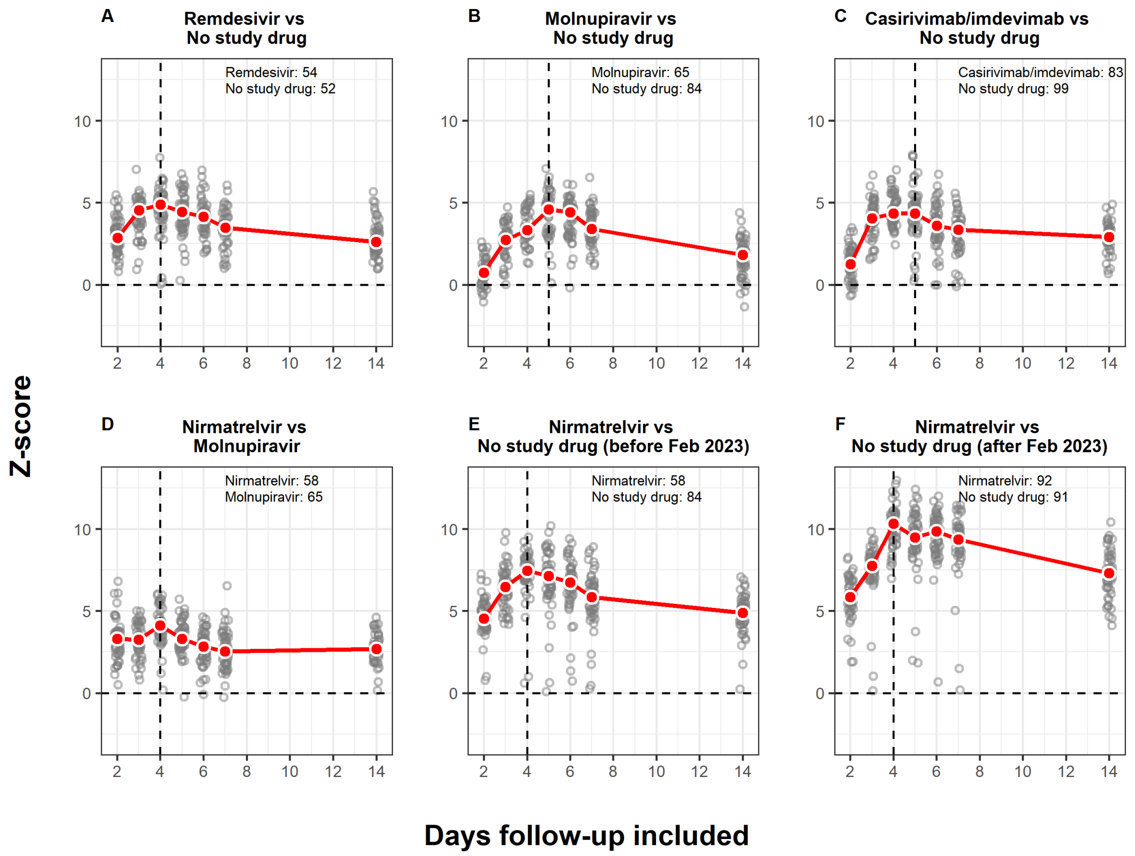

Figure S6: Z-scores for the treatment effect as a function of the follow-up duration. Grey circles represent the estimated z-score for each bootstrap iteration, while red circles (line) show the median estimates across 50 bootstrap iterations per follow-up duration. The vertical dashed line indicates the follow-up duration that maximises z-scores. This ranged between 4 and 5 days of serial sampling. The comparisons only use concurrently randomised controls. Text annotations indicate number of patients in each comparison arm.

#### S3 Casirivimab/imdevimab

The casirivivab/imdevimab resistant hypothesised subgroup is defined as: (i) G to S amino acid change in the 446 residue OR (ii) any amino acid change in the 444 residue .

#### S4 Tixagevimab/cilgavimab

The tixagevimab/cilgavimab resistant hypothesised subgroup is defined by: (i) any amino-acid change from wild-type in the 486 residue of the spike protein AND (ii) an amino-acid change in the 346 residue OR the 444 residue.
